# Body mass index and adiposity influence responses to immune checkpoint inhibition in endometrial cancer

**DOI:** 10.1101/2024.06.07.24308618

**Authors:** Nicolás Gómez-Banoy, Eduardo Ortiz, Caroline S. Jiang, Christian Dagher, Carlo Sevilla, Jeffrey Girshman, Andrew Pagano, Andrew Plodkowski, William A. Zammarrelli, Jennifer J. Mueller, Carol Aghajanian, Britta Weigelt, Vicky Makker, Paul Cohen, Juan C. Osorio

## Abstract

**Background:** Obesity is the foremost risk factor in the development of endometrial cancer (EC). However, the impact of obesity on the response to immune checkpoint inhibitors (ICI) in EC remains poorly understood. This retrospective study investigates the association between body mass index (BMI), body fat distribution, and clinical and molecular characteristics of EC patients treated with ICI.

**Methods:** We analyzed progression-free survival (PFS) and overall survival (OS) in EC patients treated with ICI, categorized by BMI, fat mass distribution, and molecular subtypes. Incidence of immune-related adverse events (irAE) after ICI was also assessed based on BMI status.

**Results:** 524 EC patients were included in the study. Overweight and obese patients exhibited a significantly prolonged PFS and OS compared to normal BMI patients after treatment with ICI. Multivariable Cox regression analysis confirmed the independent association of overweight and obesity with improved PFS and OS. Elevated visceral adipose tissue (VAT) was identified as a strong independent predictor for improved PFS to ICI. Associations between obesity and OS/PFS were particularly significant in the copy number-high/*TP53*abnormal (CN-H/*TP53*abn) EC molecular subtype. Finally, obese patients demonstrated a higher irAE rate compared to normal BMI individuals.

**Conclusion:** Obesity is associated with improved outcomes to ICI in EC patients and a higher rate of irAEs. This association is more pronounced in the CN-H/*TP53*abn EC molecular subtype.

**Funding:** NIH/NCI Cancer Center Support Grant P30CA008748 (MSK). K08CA266740 and MSK Gerstner Physician Scholars Program (J.C.O). RUCCTS Grant #UL1 TR001866 (N.G-B and C.S.J). Cycle for survival and Breast Cancer Research Foundation grants (B.W).

## Introduction

Endometrial cancer (EC) constitutes the leading cause of gynecologic cancer-related death in the United States, and one of the few cancer types with increasing incidence and disease-associated mortality (1). Obesity is one of the main drivers in the development of EC (2, 3), with a clear stepwise correlation between body mass index (BMI) and the risk of developing EC (4). Elevated body weight is also associated with worse prognosis in patients with this malignancy (5). Mechanistically, obesity induces dysfunction in the adipose tissue (AT), which has been implicated in promoting the progression and growth of EC cells (6) and triggering a dysregulated inflammatory state (7). However, there is a paucity of data regarding the influence of obesity on the response to immune-based therapies. This gap in knowledge is particularly important given that 80% of EC-diagnosed women are obese (8), and immune checkpoint inhibitors (ICI) are becoming a cornerstone for the treatment of EC (9).

Monoclonal antibodies blocking inhibitory checkpoints have recently changed the front-line treatment paradigm for advanced and recurrent EC. Dorstarlimab, a programmed cell death receptor-1 (PD-1) blocker, is now first line therapy in conjunction with chemotherapy for patients with advanced EC based on results from the RUBY trial (10). Similarly, Pembrolizumab, another PD-1 inhibitor in combination with chemotherapy, has recently shown improved progression free survival (PFS) in the frontline setting when compared to chemotherapy alone in the NRG-GY018 trial (11). In the second-line setting, ICI alone or in combination with the tyrosine kinase inhibitor (TKI) Lenvatinib are FDA-approved in recurrent EC after treatment with platinum-based chemotherapy in mismatch repair (MMR)–deficient and MMR-proficient EC, respectively (12, 13). Despite these clinical advances, there is a lack of validated clinical, molecular, and immunological biomarkers that can predict response to these therapies. To this end, one of the most intriguing findings in patients treated with ICI for non-EC malignancies is the “obesity paradox”, in which obese patients treated with ICI have improved outcomes compared to lean patients (14). Furthermore, higher BMI may also correlate with the rate of immune related adverse events (irAEs) (15), suggesting that obesity might promote disruption of immune tolerance against both tumor and normal cells. While these observations have been described in a few solid tumors (16–20), the heterogeneity across different studies and the attenuation of these associations after adjusting for relevant clinical factors underscore the need for further investigation (18, 21). Importantly, this clinical association has yet to be explored in the context of EC.

Given the high prevalence of obesity in EC and the prominence of ICI in its management, this retrospective study aims to define whether obesity influences clinical outcomes in women with EC after treatment with ICI. By characterizing clinical markers for obesity, body fat distribution and molecular EC subtypes, we found a strong association between overweight/obesity and improved clinical outcomes in EC patients treated with ICI alone or in combination with Lenvatinib. Notably, this favorable prognostic impact remained independent of clinicopathological and molecular subtyping of EC. Additionally, after assessment of body fat distribution, we found that increased visceral adipose tissue (VAT) is particularly associated with the improved clinical outcomes observed in our cohort. Finally, obesity was also linked to elevated rates of irAEs after immunotherapy. Collectively, these findings highlight the role of increased adiposity in modulating the response to ICI and their side effect profile in EC.

## Results

### Characteristics of patients with EC treated with ICI categorized by BMI

We retrospectively screened 768 patients diagnosed with EC that underwent treatment with ICI at Memorial Sloan Kettering Cancer Center (MSK) from November 2015 to November 2022. Out of these, 524 patients with recurrent, advanced, or metastatic EC were included in the final analysis (**Figure 1**). The main reason for exclusion was patients receiving ICI therapy to treat a non-EC malignancy. Underweight patients (BMI < 18.5 kg/m^2^) were also excluded from the analysis (**Figure 1**). The baseline clinical characteristics (at the start of ICI) of the patients included in the final analysis are shown in **Table 1**. Across the entire study cohort, the median age was 67 years (range 30 – 94), and the median BMI was 29.1 kg/m^2^. Most patients (85%) received anti-PD-1 therapy, while 15% received anti-PD-L1 therapy. Regarding the combination of ICI with other anticancer therapies, 307 patients (59%) were treated with pembrolizumab in combination with Lenvatinib. The majority of patients received ICI therapy as second (54%) or third line (27%) of treatment. Additionally, 437 patients (83%) underwent molecular subtyping, and 500 (95%) had a baseline computed tomography (CT) for determination of fat distribution. When categorized by BMI before the start of ICI therapy, 128 patients (24%) had a normal BMI (18.5 – 24.9 kg/m^2^), whereas 163 (31%) were overweight (BMI 25 – 29.9 kg/m^2^), and 233 (44%) were obese (BMI ≥30 kg/m^2^). Except for self-reported race and age, no significant differences in baseline characteristics were observed among the BMI groups. The number of patients with elevated subcutaneous adipose tissue (SAT), visceral adipose tissue (VAT), and VAT/SAT ratio increased from normal BMI to overweight to obese patients **(Table 1).**

**Figure 1.**
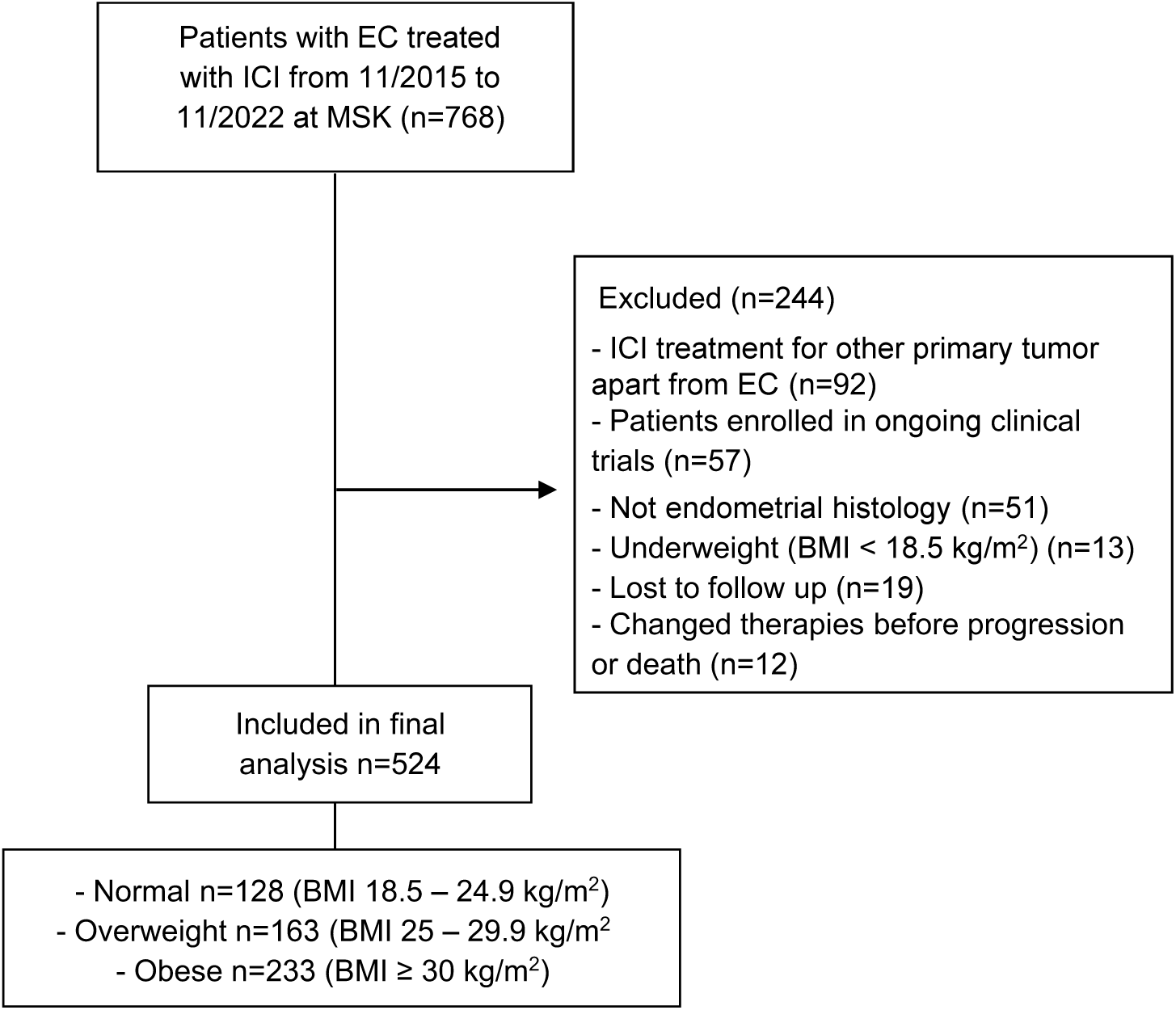
Consort diagram of the study population selection including exclusion criteria. EC, endometrial cancer; ICI, immune check-point inhibitor; MSK, Memorial Sloan Kettering Cancer Center; BMI, body mass index.

**Table 1.** Clinical characteristics of EC patients treated with ICI categorized by BMI. ^A^ Molecular subtyping was available for 437 patients. ^B^ Pre-treatment abdominal CT scans were available for 500 patients. BMI, body mass index; ECOG, Eastern Cooperative Oncology Group; CPNH/*TP53*abn, copy number-high/*TP53*abnormal; CNL/NSMP, copy number-low/no specific molecular profile; MSI-H, microsatellite instability high; VAT, visceral adipose tissue; SAT, subcutaneous adipose tissue. P values were calculated using Kruskal-Wallis, One-way ANOVA, chi-squared, or Fisher’s exact tests.

|  | All n=524 | Normal BMI<br>n=128 | Overweight<br>n=163 | Obese n=233 | p value |
| --- | --- | --- | --- | --- | --- |
| **Median age (range)** | 67 (30 - 94) | 67 (41 – 89) | 68 (43 – 94) | 66 (30 - 91) | 0.02 |
| **Median BMI, kg/m<sup>2</sup> (range)** | 29.1 (18.5 – 59.4) | 22.6 (18.5 – 24.9) | 27.3 (25 – 29.9) | 34.7 (30 – 59.4) | <0.0001 |
| **Self-reported race - No (%)** |  |  |  |  |  |
| • White: | 358 (68) | 84 (66) | 121 (74) | 153 (66) | 0.004 |
| • Black: | 77 (15) | 14 (11) | 20 (12) | 43 (18) |  |
| • Asian: | 41 (8) | 16 (13) | 15 (9) | 10 (4) |  |
| • Other: | 48 (9) | 14 (11) | 7 (4) | 27 (12) |  |
| **Histology – No (%)** |  |  |  |  |  |
| • Endometroid | 205 (39) | 44 (34) | 60 (37) | 101 (43) | 0.2 |
| o Low grade (1,2) | 131 (25) | 32 (25) | 39 (24) | 60 (26) |  |
| o High grade (3) | 74 (14) | 12 (9) | 21 (13) | 41 (18) |  |
| • Serous | 136 (26) | 27 (21) | 50 (31) | 59 (25) |  |
| • Mixed/high-grade NOS | 75 (14) | 19 (15) | 22 (13) | 34 (15) |  |
| • Carcinosarcoma | 70 (13) | 24 (19) | 20 (12) | 26 (11) |  |
| • Clear Cell | 23 (4) | 7 (5) | 8 (5) | 8 (3) |  |
| • Un- / Dedifferentiated | 15 (3) | 7 (5) | 3 (2) | 5 (2) |  |
| **Checkpoint inhibitor – No (%)** |  |  |  |  |  |
| • Pembrolizumab: | 423 (81) | 95 (74) | 131 (80) | 197 (85) | 0.15 |
| • Durvalumab: | 74 (14) | 22 (17) | 26 (16) | 26 (11) |  |
| • Nivolumab: | 17 (3) | 7 (5) | 5 (3) | 5 (2) |  |
| • Other: | 10 (2) | 4 (3) | 1 (1) | 5 (2) |  |
| **Combination therapies – No (%)** |  |  |  |  |  |
| • Lenvatinib/pembrolizumab: | 307 (59) | 71 (55) | 98 (60) | 138 (59) | 0.73 |
| • Tremelimumab/durvalumab: | 35 (7) | 11 (9) | 12 (7) | 12 (5) |  |
| • ICI alone: | 172 (33) | 42 (33) | 51 (31) | 79 (34) |  |
| • Other combination: | 10 (2) | 4 (3) | 2 (1) | 4 (2) |  |
| **ECOG Performance Status– No (%)** |  |  |  |  |  |
| • 0: | 253 (48) | 66 (52) | 91 (56) | 96 (41) | 0.06 |
| • 1: | 252 (48) | 58 (45) | 67 (41) | 127 (55) |  |
| • 2-3 | 19 (4) | 4 (3) | 5 (3) | 10 (4) |  |
| **Mean cycles per month – No (SD)** | 1.2 (± 0.5) | 1.3 (± 0.5) | 1.2 (± 0.6) | 1.1 (± 0.5) | 0.1 |
| **Stage at diagnosis (1,2 vs 3,4) – No (%)** |  |  |  |  |  |
| • 1,2: | 203 (39) | 46 (36) | 61 (37) | 96 (41) | 0.57 |
| • 3,4: | 321 (61) | 82 (64) | 102 (63) | 137 (59) |  |
| **Previous lines of therapy – No (%)** |  |  |  |  |  |
| • 0 | 31 (6) | 9 (7) | 11 (7) | 11 (5) | 0.25 |
| • 1 | 281 (54) | 56 (44) | 90 (55) | 135 (58) |  |
| • 2 | 141 (27) | 40 (31) | 42 (26) | 59 (25) |  |
| • ≥ 3 | 71 (14) | 23 (18) | 20 (12) | 28 (12) |  |
| **Previous pelvic radiotherapy – No (%)** |  |  |  |  |  |
| • Yes | 288 (55) | 65 (51) | 96 (59) | 127 (55) | 0.38 |
| • No | 236 (45) | 63 (49) | 67 (41) | 106 (45) |  |
| **Molecular subtype – No (%) <sup>A</sup>** |  |  |  |  |  |
| • CN-H/*TP53*abn | 256 (59) | 66 (58) | 77 (58) | 113 (59) | 0.06 |
| • MSI-H | 97 (22) | 21 (18) | 25 (19) | 51 (27) |  |
| • CN-L/NSMP | 81 (19) | 25 (22) | 31 (23) | 25 (13) |  |
| • POLE | 3 (0.7) | 2 (2) | 0 (0) | 1 (0.5) |  |
| **SAT – No (%) <sup>B</sup>** |  |  |  |  |  |
| • Low (≤270 cm<sup>2</sup>) | 251 (50) | 116 (93) | 102 (65) | 33 (15) | <0.0001 |
| • High (>270 cm<sup>2</sup>) | 249 (50) | 9 (7) | 54 (35) | 186 (85) |  |
| **VAT No (%) <sup>B</sup>** |  |  |  |  |  |
| • Low (≤112 cm<sup>2</sup>) | 251 (50) | 112 (90) | 98 (63) | 41 (19) | <0.0001 |
| • High (>112 cm<sup>2</sup>) | 249 (50) | 13 (10) | 58 (37) | 178 (81) |  |
| **VAT/SAT ratio <sup>B</sup> (%)** |  |  |  |  |  |
| • Low (≤0.3723) | 250 (50) | 82 (66) | 80 (51) | 88 (40) | <0.0001 |
| • High (>0.3723) | 250 (50) | 43 (34) | 76 (49) | 131 (60) |  |

### Association between BMI and clinical outcomes after treatment with ICI in patients with EC

First, we investigated whether an elevated BMI could influence the response to ICI in all EC patients included in the analysis. Survival analyses were performed after initiation of ICI therapy, revealing that patients categorized as overweight or obese exhibited a significantly prolonged PFS when compared to those with normal BMI after treatment with ICI (Overweight vs normal BMI: median 6.5 vs. 4.5 months, HR 0.71, 95% CI 0.55 – 0.93, p=0.0112. Obese vs. normal BMI: median 7.8 vs. 4.5 months, HR 0.61 95% CI 0.47 – 0.78, p<0.0001) **(Figure 2A).** Furthermore, patients with overweight and obesity demonstrated a significantly prolonged OS compared to patients with normal BMI after ICI (Overweight vs. normal BMI: median 27 vs. 15.2 months, HR 0.61, 95% CI 0.45 – 0.83, p=0.0018. Obese vs. normal BMI: median 22 vs. 15.2 months, HR 0.65 95% CI 0.49 – 0.86, p=0.0026) **(Figure 2B).**

**Figure 2.**
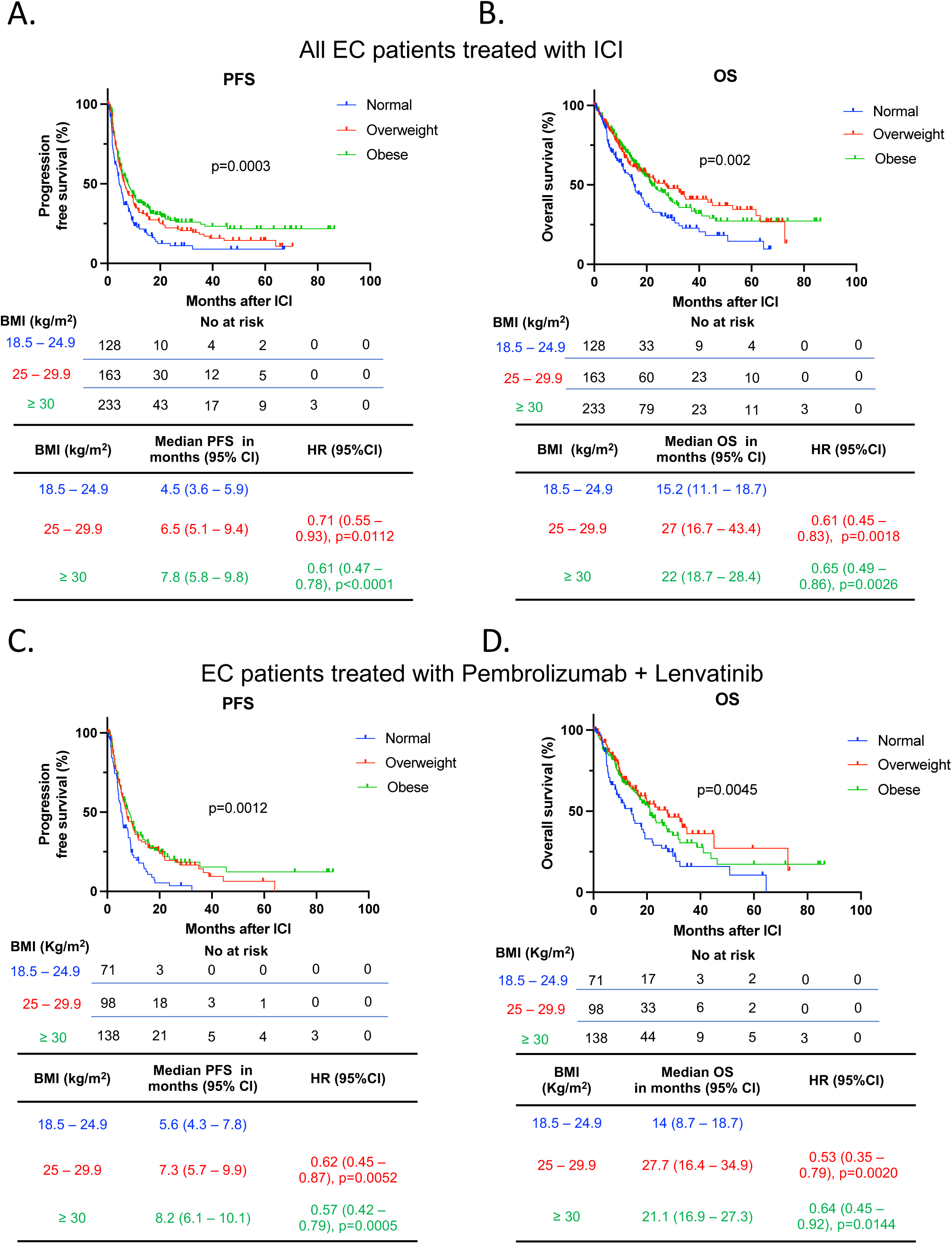
Survival outcomes of EC patients treated with ICI categorized by BMI. Kaplan-Meier curves for (A) PFS and (B) OS in patients with EC treated with ICI and categorized by BMI (Normal - BMI: 18.5 – 24.9 kg/m^2^ in blue; Overweight – BMI 25 – 29.9 kg/m^2^ in red; Obese – BMI > 30 kg/m^2^ in green) (n=524). Kaplan-Meier curves for (C) PFS and (D) OS in the subgroup of EC patients treated with the ICI pembrolizumab in combination with lenvatinib (n=307). The P values in the PFS and OS plots were calculated using a log-rank test. HRs and 95% CIs for overweight and obese patients were calculated using normal BMI as a reference. BMI, body mass index; CI, confidence interval; EC, endometrial cancer; HR, Hazard ratio; ICI, immune checkpoint inhibitor; OS, overall survival; PFS, progression free survival.

The combination of Lenvatinib with the ICI pembrolizumab is the standard of care treatment for a significant proportion of patients with MMR-proficient, advanced EC who have progressed after first-line platinum-based chemotherapy (12). As more than half of our cohort received this treatment combination **(Table 1),** we explored whether obesity was associated with clinical outcomes with this specific treatment regimen. Survival analyses in patients who received combination Lenvatinib and pembrolizumab revealed that obese and overweight patients had significantly longer PFS (Overweight vs. normal BMI: median 7.3 vs. 5.6 months, HR 0.62, 95% CI 0.45 – 0.87, p=0.0052. Obese vs. normal BMI: median 8.2 vs. 5.6 months, HR 0.57 95% CI 0.42 – 0.79, p=0.0005) and OS (Overweight vs. normal BMI: median 27.7 vs. 14 months, HR 0.53, 95% CI 0.35 – 0.79, p=0.0020. Obese vs. normal BMI median 21.1 vs. 14 months, HR 0.64 95% CI 0.45 – 0.92, p=0.0144) compared to patients with normal BMI **(Figure 2C – 2D)**.

We then explored the impact of other baseline clinical variables on the PFS and OS of EC patients after treatment with ICI therapy. Similar to BMI, univariable Cox regression analysis demonstrated that specific histological types, stage at diagnosis, number of previous lines of therapy, and molecular subtype were significantly associated with changes in PFS and OS in EC patients treated with ICI (**Tables 2 and 3**). Thus, we investigated whether BMI was independently associated with improved PFS and OS in our study cohort by controlling for these and other clinical variables. Multivariable Cox regression analysis demonstrated that baseline overweight and obese states were independently associated with improved PFS when compared to patients with normal BMI (overweight vs normal BMI: adjusted HR 0.71, 95% CI 0.54 – 0.93; obese vs normal BMI adjusted HR 0.54, 95% CI 0.42 – 0.71) **(Figure 3A)**. Similarly, overweight and obesity were independently associated with extended OS compared to normal BMI patients (Overweight vs normal BMI: adjusted HR 0.64, 95% CI 0.47 – 0.89. Obese vs normal BMI adjusted HR 0.64, CI 95% 0.48 – 0.87) (**Figure 3B**). As expected, distinct histological types (carcinosarcoma, serous, un-/dedifferentiated) and poor baseline Eastern Cooperative Oncology Group (ECOG) performance status were independent predictors of worse PFS and OS. Overall, these results suggest a paradoxical association between elevated BMI and improved responses to ICI in patients with EC, further supporting BMI as an independent predictor of clinical response to ICI.

**Figure 3.**
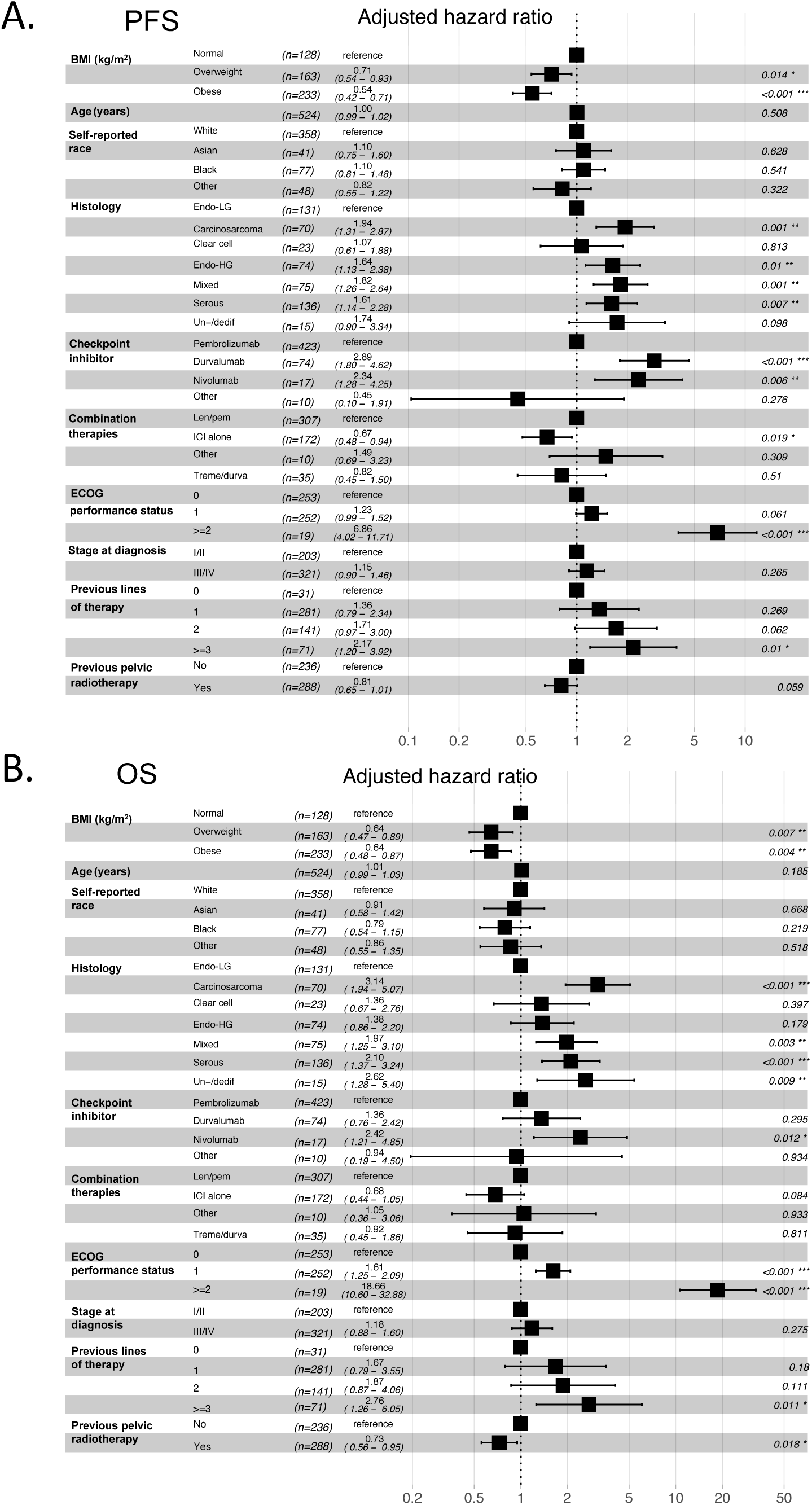
Multivariable Cox regression analysis of BMI and other clinical variables associated with response to ICI in EC patients. Forest plots of adjusted HRs and 95% CIs for patients with normal BMI (BMI 18.5 – 24.9 kg/m2) (reference group) compared to overweight (BMI 25 – 29.9 kg/m2) and obese (BMI > 30 kg/m2) for (A) PFS and (B) (OS) (n=524) Analysis was adjusted for age, self-reported race, histology, checkpoint inhibitor treatment, combination therapies, baseline performance status, stage at diagnosis, prior lines of therapy, and previous pelvic radiotherapy. BMI, body mass index; OS, overall survival; PFS, progression free survival; EC, endometrial cancer; ICI, immune checkpoint inhibitor; Endo-LG, endometrial low grade; Endo-HG, endometrial high grade; Un-/dediff, Un-/dedifferentiated; Len/pem, Lenvatinib/pembrolizumab; Treme/durva, Tremelimumab/durvalumab; HR, Hazard ratio; CI, confidence interval.

**Table 2.** Univariable Cox regression analysis for PFS in EC patients treated with ICI. ^A^Molecular subtyping n=434 patients, excluding 3 with POLE. ^B^ Pre-treatment abdominal CT scans were available for 500 patients. Reference alludes to reference group for the analysis. BMI, body mass index; ECOG, Eastern Cooperative Oncology Group; CPNH/*TP53*abn, copy number-high/*TP53*abnormal; CNL/NSMP, copy number-low/no specific molecular profile; MSI-H, microsatellite instability high; VAT, visceral adipose tissue; SAT, subcutaneous adipose tissue.

| Baseline characteristic | Variables | Univariable HR<br>(95% CI) | p value |
| --- | --- | --- | --- |
| **BMI (kg/m<sup>2</sup>)** | Normal (reference) | 1 |  |
|  | Overweight | 0.71 (0.55 – 0.93) | 0.011 |
|  | Obese | 0.61 (0.47 – 0.77) | <0.0001 |
| **Age (years)** |  | 1 (0.99 – 1.01) | 0.83 |
| **Self-reported race** | White (reference) | 1 |  |
|  | Asian | 1.14 (0.79 – 1.65) | 0.47 |
|  | Black | 1.21 (0.92 – 1.6) | 0.18 |
|  | Other | 0.83 (0.57 – 1.22) | 0.35 |
| **Histology** | Endometrioid low grade (reference) | 1 |  |
|  | Endometrioid high grade | 1.59 (1.12 – 2.26) | 0.01 |
|  | Serous | 1.88 (1.4 – 2.53) | < 0.0001 |
|  | Mixed/high-grade NOS | 2.07 (1.48 – 2.9) | < 0.0001 |
|  | Carcinosarcoma | 2.53 (1.79 – 3.6) | < 0.0001 |
|  | Clear cell | 1.46 (0.86 – 2.47) | 0.16 |
|  | Un-/dedifferentiated | 1.76 (0.93 – 3.3) | 0.08 |
| **Checkpoint inhibitor** | Pembrolizumab (reference) | 1 |  |
|  | Durvalumab | 2.26 (1.73 – 2.94) | <0.0001 |
|  | Nivolumab | 1.63 (0.95 – 2.79) | 0.08 |
|  | Other | 0.22 (0.06 – 0.88) | 0.03 |
| **Combination therapies** | Lenvatinib/pembrolizumab (reference) | 1 |  |
|  | Tremelimumab/durvalumab | 2.22 (1.54 – 3.2) | <0.0001 |
|  | ICI alone | 0.71 (0.57 – 0.9) | 0.005 |
|  | Other combination | 2.14 (1.1 – 4.17) | 0.03 |
| **ECOG Performance Status** | 0 (reference) | 1 |  |
|  | 1 | 1.24 (1 – 1.52) | 0.04 |
|  | 2-3 | 4.28 (2.62 – 6.99) | < 0.0001 |
| **Stage at diagnosis** | 1,2 (reference) | 1 |  |
|  | 3,4 | 1.28 (1.04 – 1.58) | 0.02 |
| **Previous lines of therapy** | 0 (reference) | 1 |  |
|  | 1 | 1.9 (1.14 – 3.17) | 0.013 |
|  | 2 | 2.48 (1.47 – 4.2) | 0.0007 |
|  | ≥3 | 3.41 (1.96 – 5.92) | <0.0001 |
| **Previous pelvic radiotherapy** | No (reference) | 1 |  |
|  | Yes | 0.87 (0.71 – 1.06) | 0.17 |
| **Molecular Subtype<sup>A</sup>** | MSI-H | 1 |  |
|  | CN-H/TP53abn | 3.05 (2.19 – 4.25) | < 0.0001 |
|  | CN-L/NSMP | 2.91 (1.98 – 4.27) | < 0.0001 |
| **VAT (cm<sup>2</sup>)<sup>B</sup>** | Low (≤112 cm<sup>2</sup>) (reference) | 1 |  |
|  | High (>112 cm<sup>2</sup>) | 0.69 (0.56 – 0.85) | 0.0004 |
| **SAT (cm<sup>2</sup>)<sup>B</sup>** | Low (≤270 cm<sup>2</sup>) (reference) | 1 |  |
|  | High (>270 cm<sup>2</sup>) | 0.82 (0.67 – 1.01) | 0.062 |

**Table 3.**
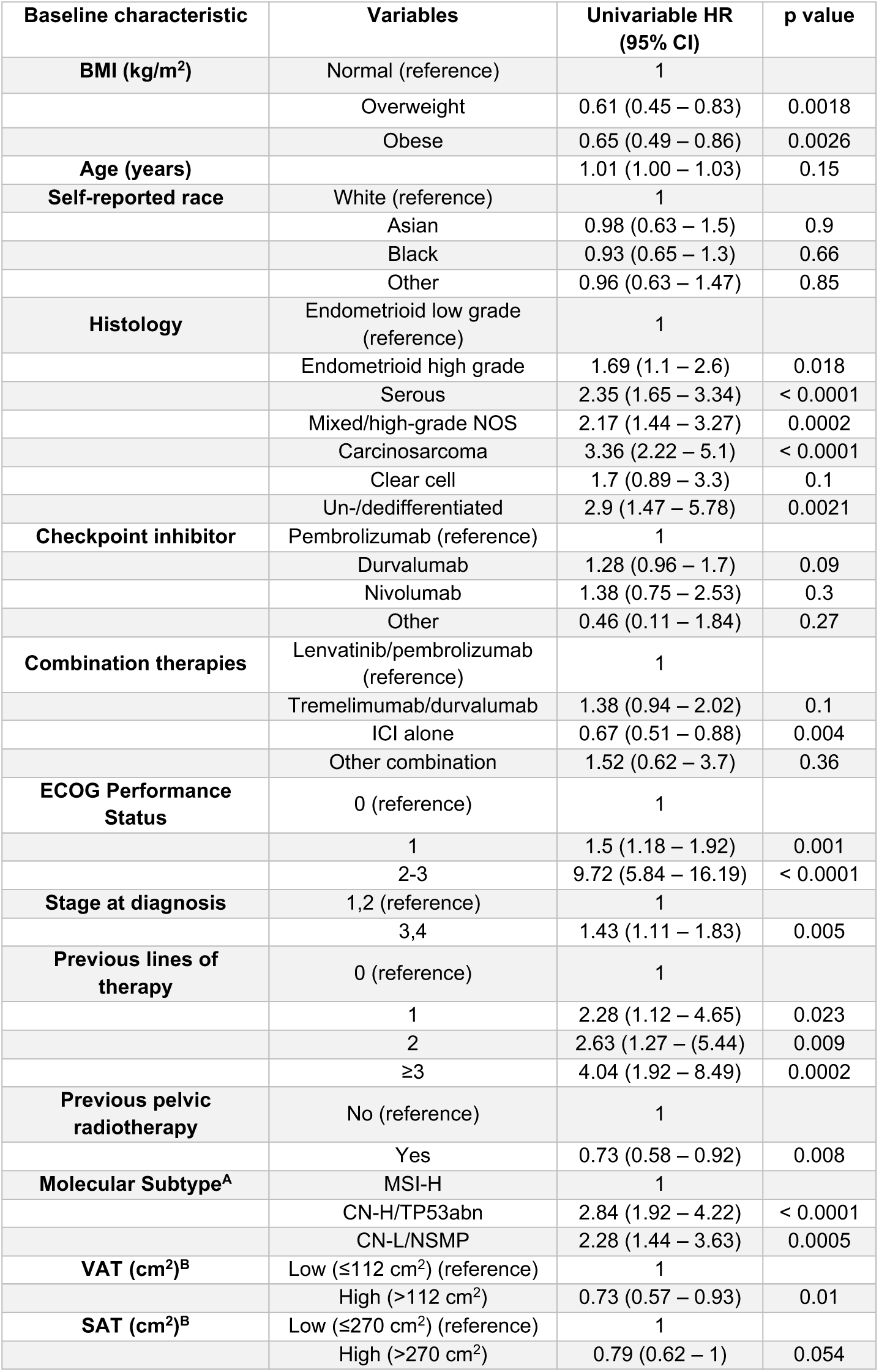
Univariable Cox regression analysis for OS in EC patients treated with ICI. ^A^Molecular subtyping n=434 patients, excluding 3 with POLE. ^B^Pre-treatment abdominal CT scans were available for 500 patients. Reference alludes to reference group for the analysis. BMI, body mass index; ECOG, Eastern Cooperative Oncology Group; CPNH/*TP53*abn, copy number-high/*TP53*abnormal; CNL/NSMP, copy number-low/no specific molecular profile; MSI-H, microsatellite instability high; VAT, visceral adipose tissue; SAT, subcutaneous adipose tissue.

### Association between fat distribution and clinical responses to ICI in patients with EC

While BMI serves as a well-established anthropometric indicator that is positively associated with cardiometabolic disease, it is important to recognize its inability to distinguish between fat and muscle mass (22). Furthermore, in the context of cancer, BMI may not precisely capture the association between AT and responses to distinct types of therapies (23). To address this limitation and assess whether specific fat distribution could predict clinical responses in patients with EC after ICI treatment, we performed two-dimensional measurements of SAT and VAT at the level of L3/L4, which have shown a strong correlation with abdominal fat volumes and cardiometabolic risk factors (24). 500 patients out of the total cohort had available baseline CT scans to assess SAT and VAT area.

BMI correlated with both SAT (r = 0.79, p < 0.0001) and VAT area (r = 0.71, p < 0.0001) **(Supplemental Figure 1)**. We then categorized EC patients based on their median VAT (112 cm^2^) or SAT (270 cm^2^) area and examined their response to ICI, as previously performed in other studies (18). In patients with high VAT area, the median PFS post-ICI was significantly prolonged compared to those with low VAT area (Median 7.8 vs. 5.4 months. HR 0.69, 95% CI 0.56 – 0.85, p = 0.0003) (**Figure 4A**). Furthermore, a high VAT area was associated with significantly prolonged OS compared to patients with low VAT area (Median 25.9 vs. 19.2 months. HR 0.73, 95% CI 0.57 – 0.93, p = 0.0096) (**Figure 4B**). In contrast, the relationship between SAT and survival outcomes was less pronounced. Among patients with EC and high SAT area, there was a numerically but not statistically significant improvement in PFS after ICI treatment compared to those with low SAT area (median 7.2 vs. 5.8 months. HR 0.82, 95% CI 0.67 – 1.01, p = 0.06) (**Figure 4C**). Similarly, an elevated SAT area was numerically associated with prolonged OS compared to EC patients with a low SAT area (median 23.1 vs. 19.5 months. HR 0.79, 95% CI 0.62 – 1, p=0.0531) (**Figure 4D**). To further characterize the association between body fat composition and clinical outcomes, we stratified VAT and SAT by quartiles. We found an incremental association between VAT area and PFS, but not OS, with patients in the highest quartile of VAT area showing a significant increase in PFS compared to patients in the lowest VAT area quartile (median 8.3 vs 5.7 months. HR 0.65, 95% CI 0.48 – 0.87, p=0.004) (**Supplemental Figure 2A-B**); in contrast, no association with PFS or OS was observed in the analysis of SAT area by quartiles **(Supplemental Figure 2C-D)**. Prior studies have suggested that the ratio between VAT and SAT could be a better predictor of cardiometabolic risk compared to VAT area measurement and BMI (25, 26). Hence, we determined the VAT/SAT ratio in our cohort and stratified patients in high and low VAT/SAT ratio according to the median (0.3723). Patients with a high VAT/SAT ratio exhibited a significant improvement in PFS (median 7.25 vs 5.5 months. HR 0.75, 95% CI 0.61 – 0.92, p=0.0049), but not OS (**Supplemental Figure 3A and 3B**). In a subgroup analysis performed in patients treated with Lenvatinib and Pembrolizumab (n=296), we observed a trend towards both high VAT and SAT being associated with improved PFS, aligning with the significant results obtained in the larger cohort (**Supplemental Figure 4A-4D**).

**Figure 4.**
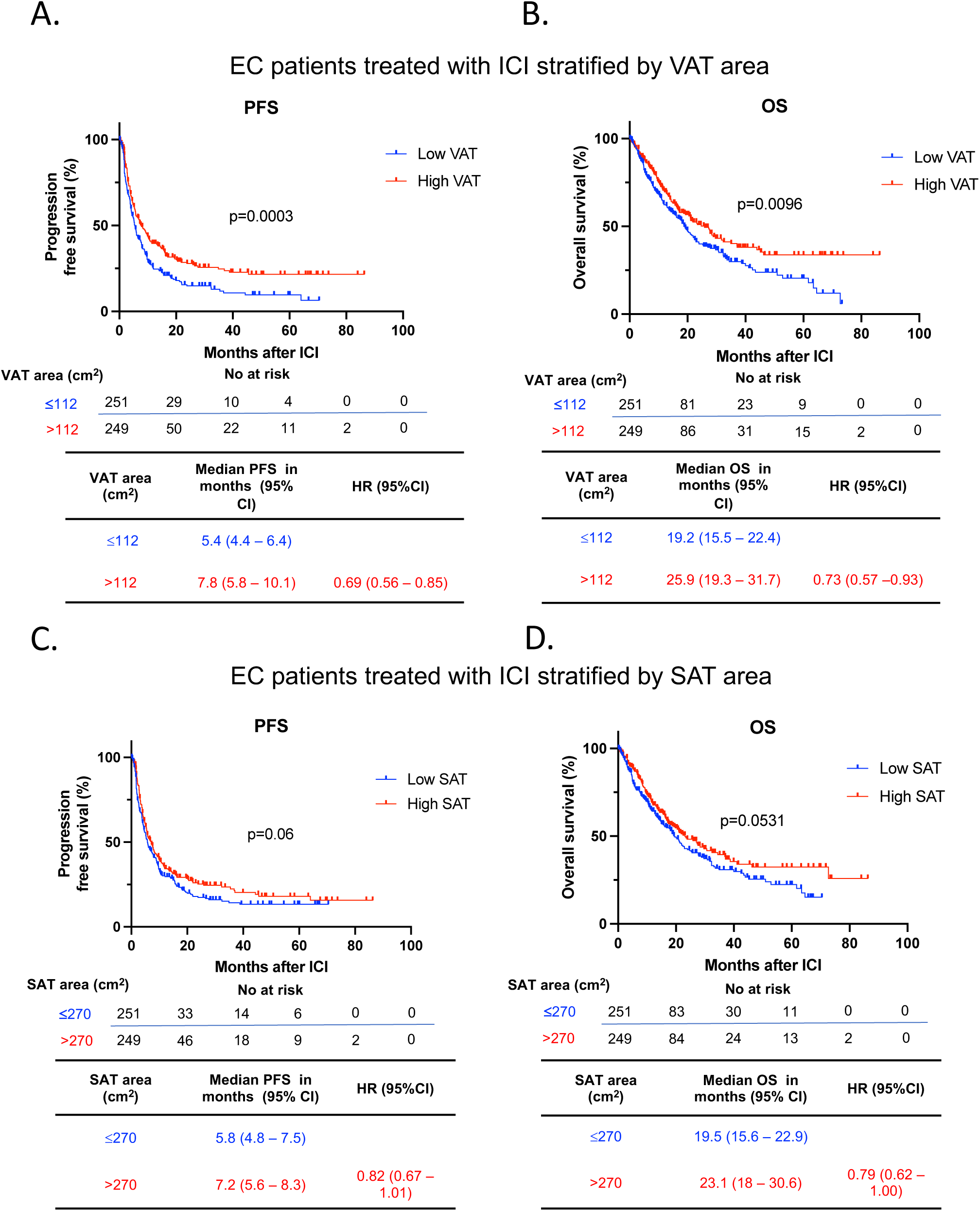
Survival outcomes after ICI in EC stratified by VAT and SAT area. Kaplan-Meier curves for (A) PFS and (B) OS in patients with EC following ICI treatment stratified by low and high VAT area (n=500) (Low VAT area: ≤112 cm^2^ in blue; High VAT area: >112 cm^2^ in red). Kaplan-Meier curves for (C) PFS and (D) OS in patients with EC following ICI treatment stratified by low and high SAT area (Low SAT area: ≤270 cm^2^ in blue; High SAT area: >270 cm^2^ in red) (n=500). Patients were categorized as low or high VAT /SAT based on the median SAT and VAT of the entire cohort. The P values in the PFS and OS plots were calculated using a log-rank test. BMI, body mass index; OS, overall survival; PFS, progression free survival; EC, endometrial cancer; ICI, immune checkpoint inhibitor; VAT, visceral adipose tissue; SAT, subcutaneous adipose tissue; HR, Hazard ratio; CI, confidence interval

To further interrogate VAT and SAT area as independent predictors for the response to ICI in EC, we performed a multivariable Cox regression analysis to control for other relevant clinical variables (**Supplemental Figure 5 and 6**). High VAT area was independently associated with improved PFS (adjusted HR 0.73, 95% CI 0.59 – 0.91) following ICI treatment (**Supplemental Figure 5A**). High SAT was also found to be independently associated with prolonged PFS (adjusted HR 0.77, 95% CI 0.621 – 0.96) although this association was less profound (**Supplemental Figure 6A**). Neither high VAT nor high SAT were associated with OS (**Supplemental Figures 5B and 6B**). Overall, these results suggest that increased VAT (and to a lesser extent SAT) in obese patients may influence the improvement in clinical responses to ICI in EC.

### Association of BMI and clinical responses after ICI across EC molecular subtypes

Of the 524 patients in the total cohort, 437 (83%) had molecular subtyping performed using an integrated molecular-immunohistochemistry approach (27). The clinical characteristics of this subgroup of patients are shown in **Supplemental Table 1**. Within this cohort, 256 (59%) ECs were classified as CN-H/*TP53*abn, 97 (22%) as MSI-H, 81 (19%) as CN-L/NSMP, and three (0.7%) as POLE (**Table 1 and Supplemental Table 1**). Akin to previous reports (27, 28), MSI-H and *POLE* patients had higher PFS and OS compared to patients with CN-H/TP53abn and CN-L/NSMP in this set of EC patients treated with ICI (**Supplemental Figures 7A and 7B**).

To determine whether BMI influenced responses to ICI across EC molecular subtypes, we built a separate multivariable Cox regression model in this subgroup accounting for molecular classification and clinicopathological features with n=434 patients, excluding the POLE molecular subtype due to the small number of patients (n=3). Overweight and obesity status remained independently associated with improved PFS (overweight vs normal BMI: adjusted HR 0.58, 95% CI 0.43 – 0.79; obese vs normal BMI: adjusted HR 0.53, 95% CI 0.4 – 0.71) and OS when compared to patients with normal BMI (overweight vs normal BMI: adjusted HR 0.5, 95% CI 0.35 – 0.72; obese vs normal BMI: adjusted HR 0.68, 95% CI 0.49 – 0.95). Additionally, ECOG performance status, specific histology types, and molecular subtype were confirmed to be independently associated with PFS and OS (**Supplemental Figure 8A and 8B**).

We then performed an exploratory subgroup analysis by molecular subtype class. In CN-H/*TP53*abn EC (n=256), obese and overweight patients had significantly prolonged PFS (Overweight vs normal BMI: median 5.8 vs. 4.0 months, HR 0.67, 95% CI 0.47 – 0.96, p=0.0264. Obese vs. normal BMI: median 6.7 vs. 4.0 months, HR 0.55, 95% CI 0.39 – 0.76, p=0.0003) and OS (Overweight vs normal BMI: median 20.9 vs. 14.3 months, HR 0.5, 95% CI 0.32 – 0.76, p=0.0012. Obese vs. normal BMI: median 21.1 vs. 14.3 months, HR 0.64, 95% CI 0.45 – 0.94, p=0.0193) when compared to normal BMI patients post-ICI (**Figure 5A and Figure 5B**). Regarding body fat distribution, amongst CN-H/ *TP53*abn EC patients with available baseline CT scan (n=249), high VAT was associated with improved PFS (median 6.86 vs 5.18 months. HR 0.68, 95% CI 0.51 – 0.89, p=0.0047) but not OS (**Supplemental Figure 9A and 9B**). High SAT was also associated with improved PFS (median 5.93 vs 5.25 months. HR 0.75, 95% CI 0.57 – 0.99, p=0.0441) and OS (19.86 vs 15.96 months. HR 0.72, 95% CI 0.52 – 0.99, p=0.0449) (**Supplemental Figure 9C and 9D**). In CN-L/NSMP EC (n=81), obese and overweight patients had a significantly prolonged PFS to ICI compared to individuals with normal BMI (Overweight vs normal BMI: median 6.5 vs. 4 months, HR 0.54, 95% CI 0.31 – 0.95, p = 0.0296. Obese vs. normal BMI: median 7.5 vs. 4 months, HR 0.51, 95% CI 0.27 – 0.94, p=0.032) (**Figure 5C**) with no differences in OS (**Figure 5D**). Body fat distribution was also assessed in CN-L/NSMP patients with available CT scan (n=79). Patients with high VAT had a trend towards improved PFS (median 7.46 vs 4.59 months. HR 0.62 95% CI 0.38 – 1.01, p=0.0525), but not OS (**Supplemental Figure 10A and 10B**). High SAT was not associated with either improved PFS or OS (**Supplemental Figure 10C and 10D**). Finally, no differences in PFS or OS were observed in MSI-H EC across BMI categories (n=97) or VAT/SAT area categories (n=90) (**Figure 5E and 5F and Supplemental Figure 11A-11D**). Overall, our data underscores the impact of obesity and overweight on prognosis, independent of clinicopathological and molecular factors. Moreover, our analyses suggest that these relationships are particularly profound in patients with CN-H/*TP53*abn EC.

**Figure 5.**
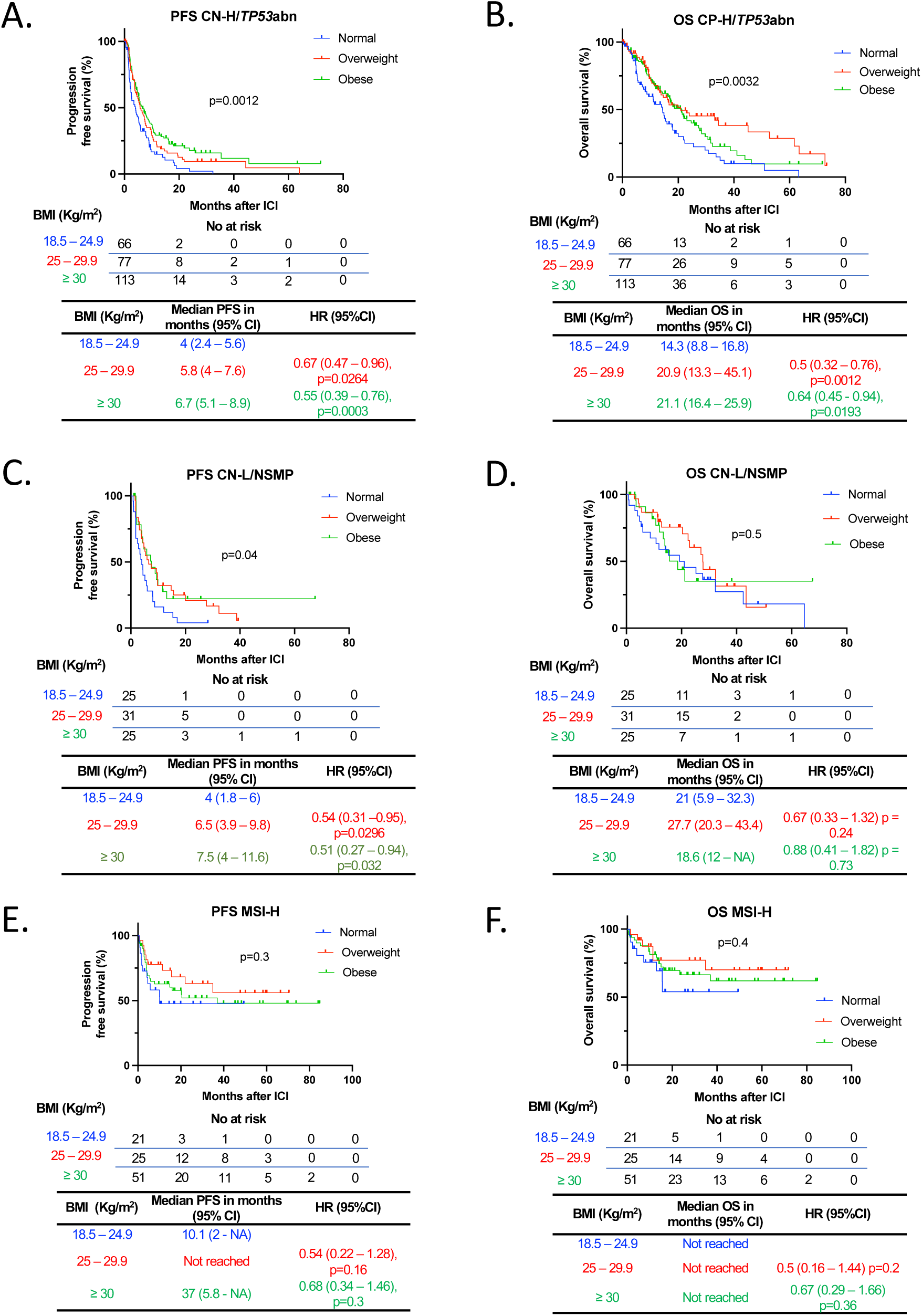
Survival outcomes following ICI in EC patients stratified by BMI across different molecular subtypes. Kaplan-Meier curves for (A) PFS and (B) OS in patients with CN-H/TP53abn EC following ICI treatment stratified by BMI. (Normal - BMI: 18.5 – 24.9 kg/m2 in blue; Overweight – BMI 25 – 29.9 kg/m2 in red; Obese – BMI > 30 kg/m2 in green) (n=256). Kaplan-Meier curves for (C) PFS and (D) OS in patients with CN-L/NSMP EC following ICI treatment stratified by BMI (n=81). Kaplan-Meir curves for (E) PFS and (F) OS in patients with MSI-H EC following ICI treatment stratified by BMI (n=97). The P values in the OS plots were calculated using a log-rank test. HRs and 95% CIs for overweight and obese patients were calculated using normal weight as a reference. BMI, body mass index; OS, overall survival; PFS, progression free survival; EC, endometrial cancer; ICI, immune checkpoint inhibitor; CN-H/TP53abnl, copy number-high/TP53abnormal; CN-L/NSMP, copy number-low/no specific molecular profile; MSI-H, microsatellite instability-high; HR, Hazard ratio; CI, confidence interval.

### Association between BMI and immune related adverse events (irAEs) in EC patients treated with ICI

irAEs are autoimmune conditions affecting any organ in the body post-ICI administration, with heterogeneous clinical presentations and poorly understood underlying biology (29). Previous studies suggest a positive association between improved clinical responses to ICI and development of irAEs (30–33). We investigated whether BMI is associated with the frequency of irAEs (assessed by Common Terminology Criteria for Adverse Events version 5) after ICI treatment. In the total cohort, the rate of irAEs of any grade was 49.6%. BMI category was significantly associated with the incidence of iRAEs (p=0.018) (**Figure 6A**). More specifically, obesity, but not overweight, was associated with increased odds of developing irAEs after ICI therapy (overweight vs normal BMI: OR 1.46, 95% CI 0.91 – 2.33. Obese vs normal BMI: OR 1.87, 95% CI 1.21 – 2.91) (**Figure 6B**). We also analyzed the incidence of irAEs in patients with high vs low VAT and SAT and did not find significant differences (**Supplemental Figure 12A and 12B**). To further characterize the link between BMI and irAEs, we stratified irAEs based on their severity (assessed by CTCAE criteria version 5) and analyzed whether BMI, VAT area or SAT area were positively associated with severe adverse events. No significant differences were found in the proportion of mild/moderate (G1/G2) vs severe (G3/G4/G5) irAEs when stratified by BMI category or high/low VAT and SAT (**Supplemental Figure 12C-E**). There was a trend towards an association between severe irAEs and BMI categories (p=0.0523).

**Figure 6.**
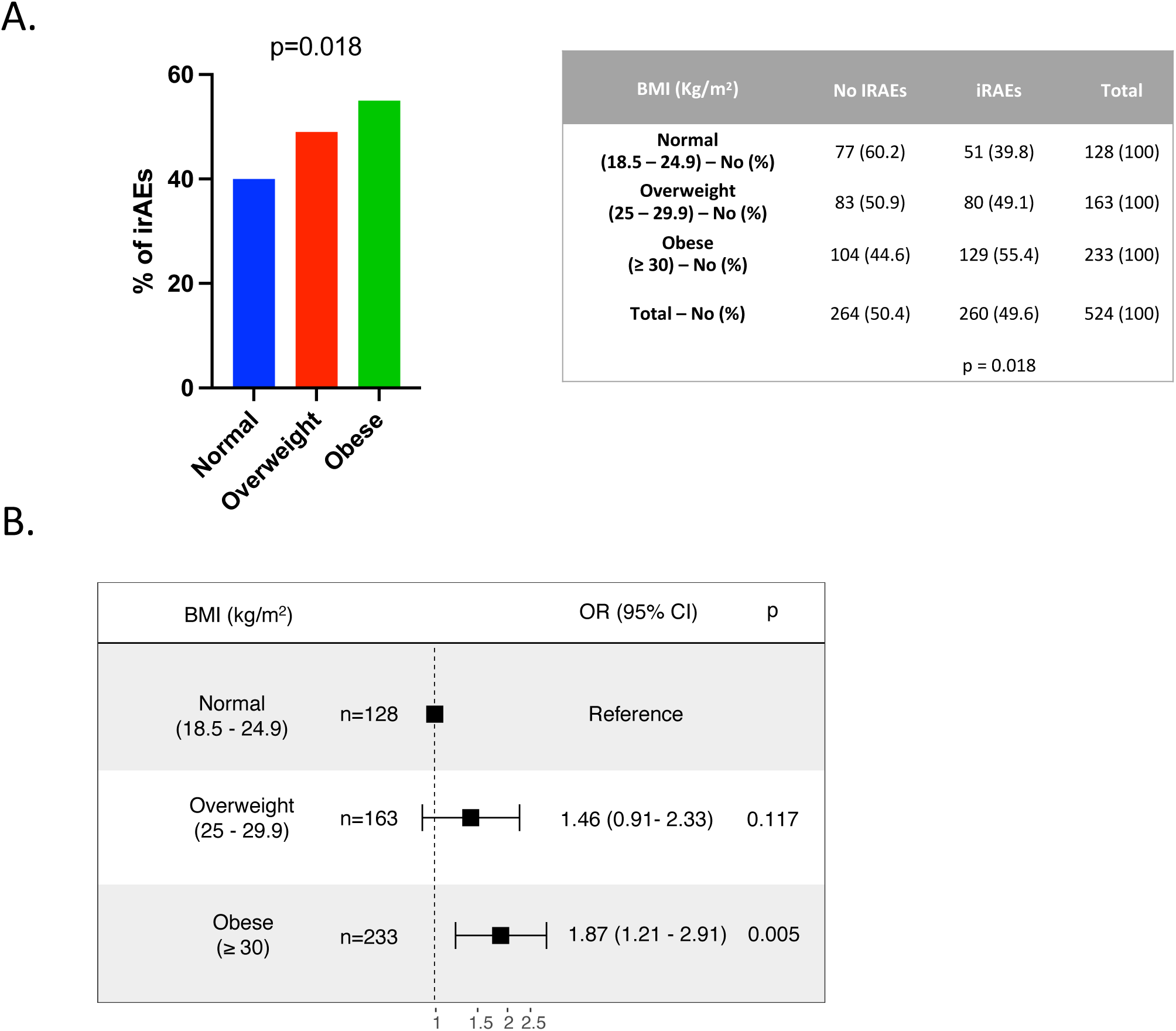
Incidence of irAEs in EC patients after treatment with ICI stratified by BMI. (A) Percentage and absolute number of irAEs across BMI categories (Normal - BMI: 18.5 – 24.9 kg/m^2^ in blue; Overweight – BMI 25 – 29.9 kg/m^2^ in red; Obese – BMI > 30 kg/m^2^ in green). Representative figure (Left) and table (right) are shown. P value in the bar graph and table was calculated using chi-squared test. (B) Forest plot of ORs and 95% CIs for patients with normal BMI (reference) compared to overweight and obese patients and their incidence of irAEs. BMI, body mass index; EC, endometrial cancer; ICI, immune checkpoint inhibitor; iRAEs, immune related adverse events; OR, odds ratio.

We then interrogated whether BMI influenced the incidence of distinct irAEs. In the whole cohort, thyroid irAEs were the most reported events (34% hypothyroidism and 14% hyperthyroidism) (**Table 4**). Next in prevalence were gastrointestinal (colitis, hepatitis, pancreatitis) (11%), skin (6%), and rheumatoid (2%) irAEs (**Table 4**). Other organ systems had less than 10 cases reported for the whole cohort (**Supplemental Table 2**). When stratified by BMI, obese patients had a numerically higher rate of hypothyroidism compared to those with normal BMI (normal BMI: 27%; overweight: 33%; obese 39%; p = 0.1) (**Table 4**); no differences in other irAE were observed across BMI categories.

**Table 4.** irAEs per organ system in EC patients after treatment with ICI stratified by BMI. Absolute number and percentage of irAEs per organ system across BMI categories Normal - BMI: 18.5 – 24.9 kg/m^2^; Overweight – BMI 25 – 29.9 kg/m2; Obese – BMI > 30 kg/m^2^. P-values were calculated with chi-squared or Fisher’s exact test. BMI, body mass index; EC, endometrial cancer; ICI, immune checkpoint inhibitor; irAEs, immune related adverse events.

| BMI (Kg/m<sup>2</sup>) | Hypo<br>thyroidism | Hyper<br>thyroidism | Skin | Colitis | Hepatitis/<br>pancreatitis | Rheumatoid | Other<br>endocrine | Nephritis |
| --- | --- | --- | --- | --- | --- | --- | --- | --- |
| **Normal**<br>**(18.5 – 24.9) - No (%)** | 35 (27) | 13 (10) | 7 (5) | 9 (7) | 5 (4) | 1 (0.8) | 1 (0.8) | 1 (0.8) |
| **Overweight**<br>**(25 – 29.9) – No (%)** | 53 (33) | 20 (12) | 9 (6) | 13 (8) | 6 (4) | 4 (2) | 5 (3) | 3 (2) |
| **Obese**<br>**(≥ 30) – No (%)** | 90 (39) | 41 (18) | 17 (7) | 16 (7) | 10 (4) | 8 (3) | 6 (3) | 4 (2) |
| **Total - No (%)** | 179 (34) | 74 (14) | 33 (6) | 38 (7) | 21 (4) | 13 (2) | 12 (2) | 8 (2) |
| **p value** | 0.1 | 0.1 | 0.7 | 0.9 | 1 | 0.3 | 0.5 | 0.8 |

### Exploratory analysis of baseline circulating white blood cells in EC patients treated with ICI

To investigate the potential mechanism behind the protective effect of overweight and obesity in patients with EC treated with ICI, we performed an exploratory analysis using the baseline levels of circulating white blood cells (WBC) as a proxy for systemic inflammation. All the patients in our cohort (n=524) had a baseline WBC count and neutrophil count (before ICI treatment), whereas 451 had baseline lymphocyte counts. We found that there were no differences between number of WBCs and neutrophils across BMI categories (**Figure 7A and 7B**). In contrast, we found that there was a higher number of absolute lymphocytes in overweight and obese patients with EC before ICI treatment (**Figure 7C**). We then calculated the neutrophil-to-lymphocyte ratio (NLR), which has been proposed as a surrogate marker of inflammation status and adaptive immune surveillance (34). Furthermore, low NLR has been associated with improved outcomes to ICI in pan-cancer cohorts (34). There was a significant difference in NLR across BMI categories (p=0.0339); overweight patients had a significantly lower NLR compared to normal BMI (p=0.0118), with no differences found between obese and normal BMI categories (**Figure 7D**). These data point towards a potential role of circulating immune cells in mediating the association between elevated BMI and improved clinical outcomes in EC patients after ICI therapy.

**Figure 7.**
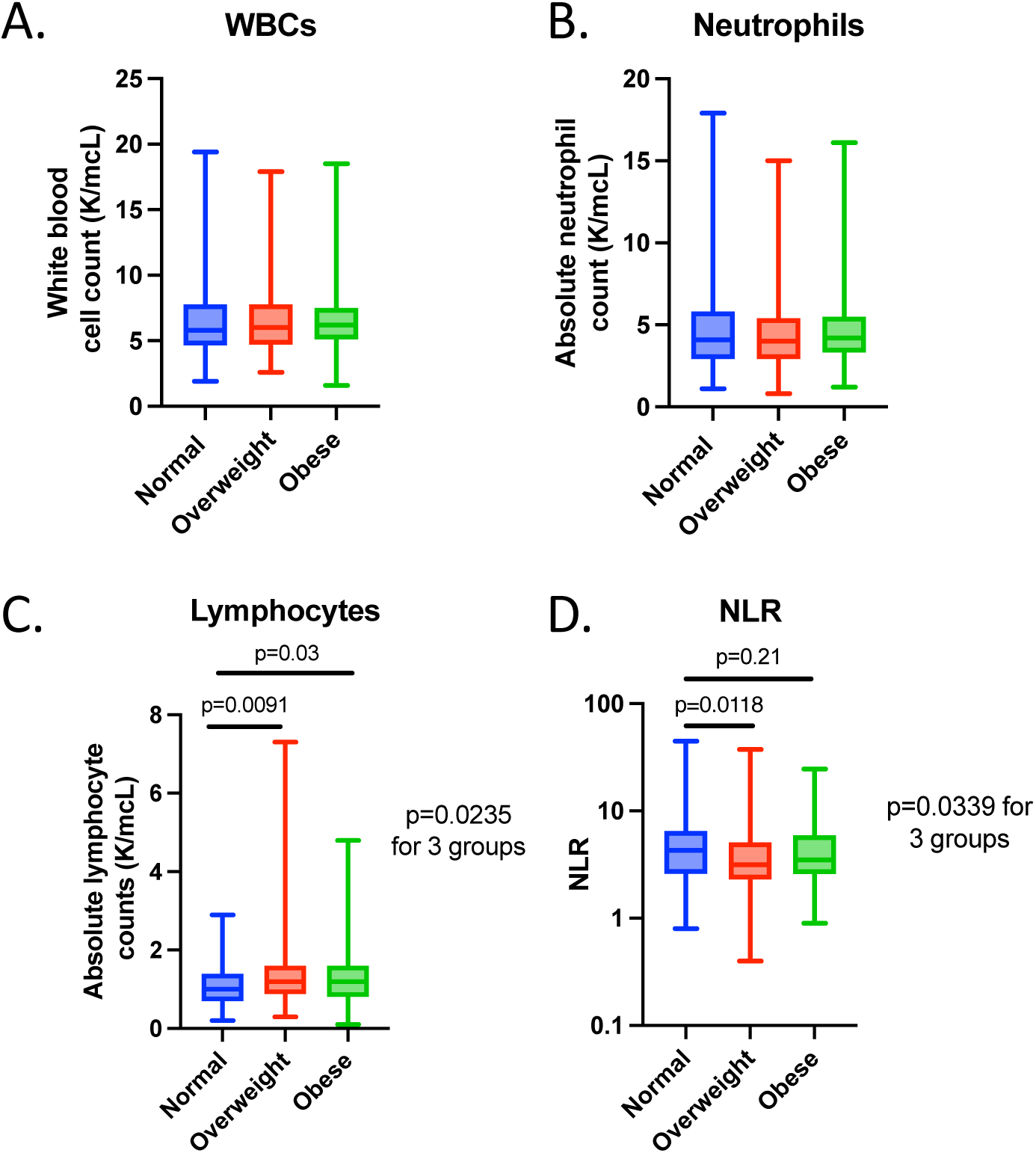
WBC, neutrophil and lymphocyte counts in EC patients treated with ICI stratified by BMI. Number of (A) WBC (B) neutrophils, (C) lymphocytes and (D) calculated NLR across BMI categories (Normal - BMI: 18.5 – 24.9 kg/m2 in blue; Overweight – BMI 25 – 29.9 kg/m2 in red; Obese – BMI > 30 kg/m2 in green). P values comparing two groups were calculated using Mann-Whitney test, P values comparing three groups were calculated using Kruskal-Wallis test. BMI, body mass index; EC, endometrial cancer; ICI, immune checkpoint inhibitor; WBC, white blood cell; NLR, neutrophil-to-lymphocyte ratio.

## Discussion

In this study, we demonstrate that overweight and obese patients with EC exhibit significantly prolonged survival following treatment with ICI compared to patients with normal BMI. Importantly, these associations remained significant after adjusting for relevant clinical factors and EC molecular subtypes. Moreover, elevated adiposity, especially in the visceral compartment, independently predicts improved PFS. Importantly, molecular classification of EC highlights that the association between obesity and response to ICI is particularly pronounced in patients with the CN-H/*TP53*abn EC subtype and is absent in patients with MSI-H EC. Finally, obesity was also associated with a higher rate of irAEs after ICI in EC patients, suggesting an enhanced immune response in this setting.

The “obesity paradox” has been investigated in other cancer types after treatment with ICI (16–20), with a first study in metastatic melanoma revealing improved survival outcomes in male obese patients receiving ICI or targeted therapy, but not in patients receiving chemotherapy (16). While this association persisted after adjusting for other clinical factors, the findings were limited to BMI categories, and other markers for obesity in this cohort were not explored. A separate study found a positive correlation between BMI and response to atezolizumab in non-small cell lung cancer (20), but no correlation was seen with the development of irAEs. In contrast, a subsequent study in patients with renal cell carcinoma showed no association between obesity and response to ICI after adjusting for other clinical variables (18). Collectively, these results suggest that obesity may have a different effect on responses to ICI depending on the type of malignancy and underscore the need for tumor-specific studies to better understand these interactions.

In line with our results, a pan-cancer study indicated that obesity and overweight status were associated with improved PFS and OS after ICI therapy, with a suggestive trend in a small subgroup of EC patients (19). Our study expands on these observations and uniquely establishes the positive correlation of obesity and elevated adiposity in EC patients with improved responses to ICI. After adjusting for multiple factors, including tumor molecular subtyping, obesity remained a significant predictor of improved clinical outcomes. Furthermore, our analyses included assessment of various body composition parameters beyond BMI, revealing a significant association between elevated VAT and favorable outcomes. Importantly, our cohort is racially and ethnically diverse, which makes our findings applicable to the real-world setting.

The mechanisms underlying the effects of obesity and adipose tissue dysfunction on immune responses during ICI treatment, remain largely underexplored. In preclinical models for melanoma, lung, colorectal and breast cancer, obese mice exhibit accelerated tumor growth and progression when compared to lean mice (35–37). These effects are partially attributed to an exhausted PD-1^high^ CD8^+^ T cell phenotype or a general decrease in CD8 T cell infiltration (35–37). Interestingly, responses to PD-1 blockade were different across these tumor models. In melanoma and lung cancer models, PD-1 blockade reinvigorated PD-1^high^ CD8^+^ T cells, resulting in enhanced antitumor activity in obese but not in lean mice (35). Of note, this T cell exhausted phenotype was partially mediated by leptin, highlighting a potential crosstalk between AT and immune responses to cancer. Conversely, PD-1 blockade did not confer additional benefit in obese mice with colorectal or breast cancer (36). Additional correlative studies in human endometrial tumor samples showed that CD8^+^ T cells and PD-L1 expression was decreased in the tumor microenvironment of patients with elevated BMI. However, PD-1, the main marker for T cell exhaustion, was not measured directly in this study. We hypothesize that obesity in EC may induce a dysfunctional CD8^+^ T cell phenotype with elevated expression of PD-1 and other inhibitory immune checkpoints. As a result, this exhausted phenotype might be more responsive to “re-invigoration” by anti-PD-1 therapy and other immunotherapies. Further prospective studies analyzing PD-1 expression in T cells from EC tumor microenvironment are warranted to confirm this hypothesis. To this end, we did find an increased number of circulating lymphocytes and a lower systemic NLR in patients with overweight and obesity in our study, highlighting a role for potential circulating immune cells in mediating this “obesity paradox”. Of note, about half of the patients in our study received treatment with the combination of the PD-1 blocker Pembrolizumab and the TKI Lenvatinib, raising the question whether this combination treatment could have a unique effect over immune responses in the context of obesity and EC. Studies analyzing the EC tumor microenvironment before and after ICI therapy and its association with circulating inflammatory factors and AT inflammation are crucial to fully dissect these mechanisms.

Our study also highlights the association between fat mass, specifically VAT, and enhanced responses to ICI therapy. These findings contrast with previous studies that identified elevated VAT as an adverse prognostic factor in patients with EC (38, 39). Transcriptomic analysis of omental VAT from women with EC revealed that patients with higher AT inflammation exhibited increased expression of genes associated with proinflammatory pathways, which may result in increased susceptibility to ICI (40). Overall, elevated body weight and AT inflammation seem to contribute to a dysfunctional immune response in EC, promoting cancer growth. Paradoxically, we hypothesize that this dysregulated immune state might confer susceptibility to ICI therapy, resulting in a protective effect in patients with obesity and increased visceral adiposity.

EC is a clinically, histologically, and molecularly heterogeneous disease. The EC molecular classification holds prognostic value (41, 42), and in certain instances, it offers predictive value into specific cancer therapies (43–45). Our study reveals that baseline BMI is a predictor of response to ICI independent of the molecular classification. Notably, in a subgroup analysis, patients with CN-H/*TP53*abn EC display a particularly strong association between elevated BMI and improved ICI outcomes. This is relevant as patients with this molecular subtype have the worst clinical outcomes (28), emphasizing the unmet need for biomarkers predicting clinical responses in this group. Furthermore, evidence suggests differences in the immune microenvironment across different EC molecular subtypes (46–48). For instance, the *TP53* mutant subtype exhibits the highest densities of both PD-1^+^ T cells and PD-L1^+^ macrophages compared to other molecular subtypes (47). Understanding how genetic alterations in EC shape the tumor immune microenvironment and influence therapy responses represents a critical knowledge gap. Prospective studies investigating these relationships across different molecular subtypes are essential to validate our findings.

Finally, our studies reveal an association between obesity and higher rates of irAEs. Most of the adverse events reported were thyroid-immune-related, likely linked to the prevalent use of Lenvatinib in our cohort (12). Of note, we found a trend towards increasing incidence of severe irAEs with BMI categories; mechanistically, it remains unclear whether the higher responses to ICI in patients with elevated BMI contribute to the higher incidence of mild/moderate irAEs. Increased T cell activation and proliferation in response to ICI, secretion of systemic pro-inflammatory cytokines, and cross reactivity in tumoral antigenicity have been suggested as potential mediators of these irAEs (49).

Our study has several limitations. First, its retrospective design highlights the need for prospective studies for further validation. However, we controlled for multiple clinical confounders, including molecular subtyping, which solidifies our findings. Second, not all the patients had available molecular characterization and baseline CT scans for body composition assessment, yet subset analysis on corresponding patients yielded results consistent with the total cohort. Finally, baseline BMI may not fully reflect weight dynamics in EC patients before and after ICI treatment, leading to potential bias in our analysis (50). To address this, we chose to complement our analyses with alternative body composition measurements, which aligned with the results from the BMI analysis.

In conclusion, our study presents clinical responses to ICI from a large cohort of EC patients stratified by BMI. Obesity and overweight were independently associated with improved survival after ICI, particularly in high-risk molecular subtypes of EC. Visceral fat mass, notably, is predominantly associated with these improved clinical responses, suggesting a potentially unique role in mediating effective immune responses in EC. Overall, our findings underscore the need for further mechanistic studies using EC biospecimen analysis and relevant EC preclinical models.

## METHODS

### Cohort characteristics

To screen for eligible patients, we extracted available electronic health record data from all patients with a histological diagnosis of EC that received treatment with ICI at Memorial Sloan Kettering Cancer Center (MSK) from November 2015 to November 2022 (n=768). We included patients that received at least 1 dose of ICI and had advanced, recurrent, or metastatic EC. We then excluded patients based on the criteria outlined in Figure 1 as follows: patients that received ICI therapy to target a primary tumor different from EC, those with non-EC histology, underweight patients as defined by BMI < 18.5, patients who received one dose of ICI and were subsequently lost to follow up (changed providers from MSK to another health institution), patients that received other anti-cancer therapy after ICI had been started before evidence of progression or death, and patients enrolled in ongoing clinical trials. Baseline patient characteristics (before ICI treatment) including age, BMI, self-reported race, previous lines of therapy, ECOG performance status, were obtained by manual chart review and used for subsequent analysis. BMI was categorized according to World Health Organization criteria as normal (18.5 – 24.9 kg/m^2^), overweight (25 – 29.9 kg/m^2^), and obese (≥ 30 kg/m^2^). Stage at diagnosis was defined by FIGO 2009 classification. Given that the combination of pembrolizumab and Lenvatinib was the most common treatment regime in our cohort, we performed a subgroup analysis of survival outcomes in these patients (n=307).

We additionally performed survival analysis stratified by adipose tissue area in patients with available baseline CT scans (a maximum of 3 months before ICI initiation) (n=5 00). Those patients with no available baseline CT scan were excluded in this subgroup analysis.

For survival analysis stratified by molecular subtype and BMI, we analyzed the subgroup of patients with these data available (n=437). All patients provided written informed consent for tumor genomic sequencing.

### Outcomes

Available clinical records were reviewed for the primary study outcomes. PFS was defined as the time from first ICI infusion to disease progression or any cause of death; patients without progression were censored at date of last office visit. OS was defined as the time from first ICI infusion to any cause of death; patients that did not die were censored at date of last office visit. Progression was assessed using the Response Evaluation Criteria in Solid Tumors (RECIST), version 1.1 (51). When formal RECIST evaluation was not available (n=365, 68%), we manually reviewed physician’s notes and imaging reports to classify overall best response using the same criteria. For consistency, all patients were reviewed by the same investigator and supervised by a senior author.

irAEs were defined according to the Common Terminology Criteria for Adverse Events (CTCAE) version 5 by manual review of the chart. Thyroid related adverse events were the most common in the cohort and were further divided into hypo and hyperthyroidism. irAEs included specific ones such as colitis, pneumonitis, hepatitis, pancreatitis, nephritis, and myocarditis. Grouped irAEs were skin (maculopapular eruptions, dermatitis, pruritus), rheumatoid (arthritis, myositis, polymyalgia rheumatica), other endocrine (diabetes, hypophysitis, adrenal insufficiency), neurological (encephalitis, meningitis), ocular (uveitis, optic neuritis), and hematologic (hemolytic anemia).

### Measurement of body fat distribution

Body fat composition variables were assessed using commercially available software (Aquarius iNtuition version 4.4.13.P6.; Tera Recon, Foster City, CA, USA). Outer abdominal circumference, subcutaneous (SAT) and visceral (VAT) adipose tissue areas (CT density range: −195-45 Hounsfield units) were semiautomatically calculated from pretreatment CT scans, using the axial plane at the L3/L4 intervertebral level (24). In cases with incorrect delimitation of the subcutaneous/ visceral fat; the radiologist manually fixed its limits using visual assessment and recalculated these variables. Based on the median of SAT area (270 cm^2^), patients were further categorized as low SAT (**≤**270 cm^2^) and high SAT area (> 270 cm^2^). Based on the median VAT area (112 cm^2^) patients were further categorized on low VAT area: **≤**112 cm^2^ and high VAT area: >112 cm^2^. Patients were additionally categorized in quartiles based on SAT and VAT area. For SAT, quartile 1 was 3 – 189 cm^2^, quartile 2 was >189 – 270 cm^2^, quartile 3 was >270 – 380 cm, and quartile 4 was >380 – 866 cm^2^. For VAT, quartile 1 was 4.5 – 56.1 cm^2^, quartile 2 was >56.1 – 112 cm^2^, quartile 3 was >112 – 172 cm^2^, and quartile 4 was >172 – 470 cm^2^ in purple. When developing sub-group analyses, high VAT and SAT areas were based on the median for each particular subgroup.

### Clinicopathologic features

Pathology reports authored by departmental gynecologic pathologists throughout the study timeframe were reviewed. These contain histopathologic data evaluated through a uniform diagnostic approach with biweekly diagnostic consensus conferences, as previously described (52). Histologic type, FIGO 2009 stage, and endometrioid tumor grade were recorded based on the patient’s initial pathologic diagnosis as previously described (27). All histologic subtypes were included (i.e., endometrioid, serous, clear cell, carcinosarcoma, un-/dedifferentiated, and mixed/high-grade not otherwise specified (NOS). The highest histologic grade for endometrioid type ECs was recorded from either the pre-operative biopsy or hysterectomy specimen.

### Molecular subtype classification

Molecular subtype using an integrated molecular-immunohistochemistry approach was performed as previously described (27). In brief, for cases with a minimum tumor purity of 20%. I) POLE molecular subtype was defined by the presence of a known POLE hotspot exonuclease domain mutation (53), ii) MSI-H molecular subtype was assigned if the MSIsensor score was ≥10 (54) and/or if MMR-deficient (MMRd) based on immunohistochemistry (IHC) MLH1, MSH2, and/or PMS2 and MSH6, iii) CN-H/*TP53*abnormal molecular subtype was assigned based on the presence of a TP53 homozygous deletion or a pathogenic driver mutation, and iv) CN-L/NSMP molecular subtype was assigned if a tumor sample did not harbor any of the defining features of the other three subtypes.

### White blood cell quantification

Absolute WBC, neutrophil and lymphocyte numbers were gathered from complete blood counts collected up to 4 weeks prior to ICI treatment. NLR was calculated as the absolute count of neutrophils divided by the absolute count of lymphocytes.

### Statistics

Kruskal-Wallis test was used to compare continuous variables across three BMI categories and Mann-Whitney test for comparing two groups. Chi-squared or Fisher’s exact test was used for categorical variables. PFS and OS curves were plotted using Kaplan-Meier method and log-rank test was used to compare survival distributions. Hazard ratios (HR) were estimated using Cox proportional hazards model. Multivariable Cox regression models included BMI category, VAT or SAT group and clinically relevant variables as covariates. Odds ratios (OR) were calculated using logistic regression. HR and OR estimates are reported with 95% confidence intervals and corresponding p-values. Spearman correlation coefficient was used to assess the linear relationship between continuous BMI and VAT or SAT area. All statistical analyses were performed using SAS Studio 3.81 and R 4.0.4. P-values<0.05 were considered statistically significant.

### Study approval

The institutional review boards at MSK approved this retrospective study.

## Data Availability

All data produced in the present study are available upon reasonable request to the authors

## Data availability

All data generated in this study are included in the article or supplementary material and are available from the corresponding authors upon further reasonable request.

## Author contributions

NGB and JCO conceived and designed the study. NGB, WAZ, CS and CD collected the clinical data. EO, JG, AP, and APl, performed the radiological analysis. NGB and CSJ conducted the statistical analysis. JCO, PC, JM, CA, BW, and VM supervised data collection, reviewed, and revised the manuscript. PC and JCO supervised the study. CD, BW, and JM performed the molecular subtype analysis. NGB and JCO wrote the original draft. All authors contributed to manuscript preparation.

## Acknowledgements

This study was supported by NIH/NCI Cancer Center Support Grant P30CA008748 (MSK). K08CA266740 and MSK Gerstner Physician Scholars Program to J.C.O. RUCCTS Grant #UL1 TR001866 to N.G-B. and C.S.J. Cycle for survival and Breast Cancer Research Foundation grants to B.W.

---

**Supplemental Figure 1.**
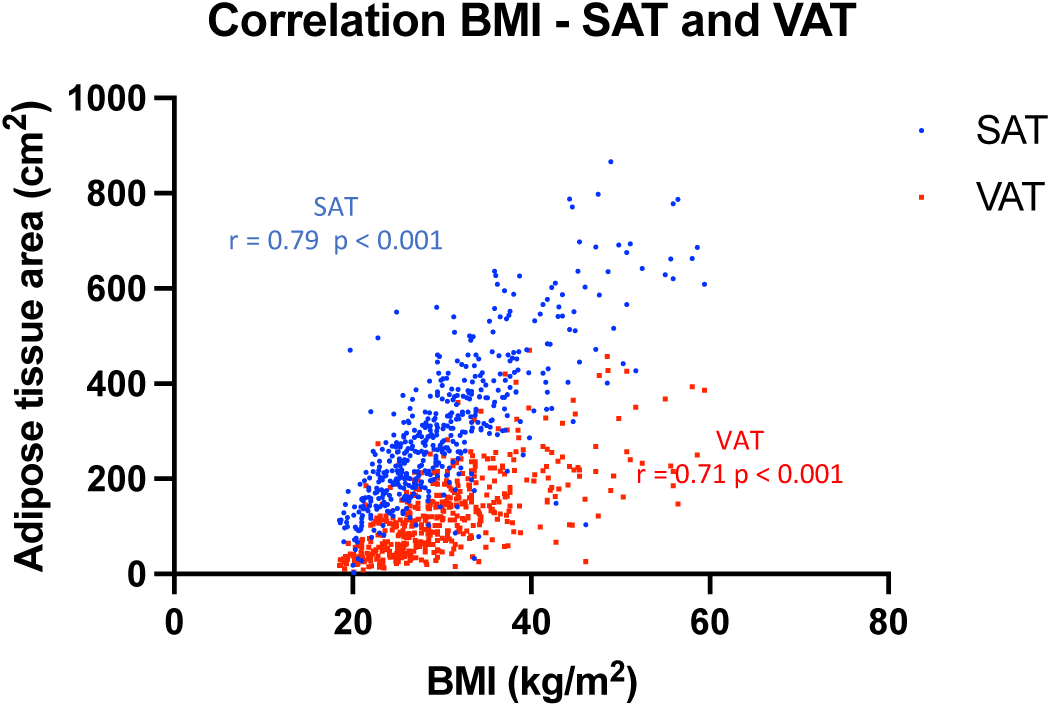
Correlation between BMI and VAT or SAT area. Scatterplots depicting the correlation between BMI and VAT and SAT area. Correlation was assessed by Spearman’s rank correlation coefficient n=500. BMI, body mass index; VAT, visceral adipose tissue; SAT, subcutaneous adipose tissue.

**Supplemental Figure 2.**
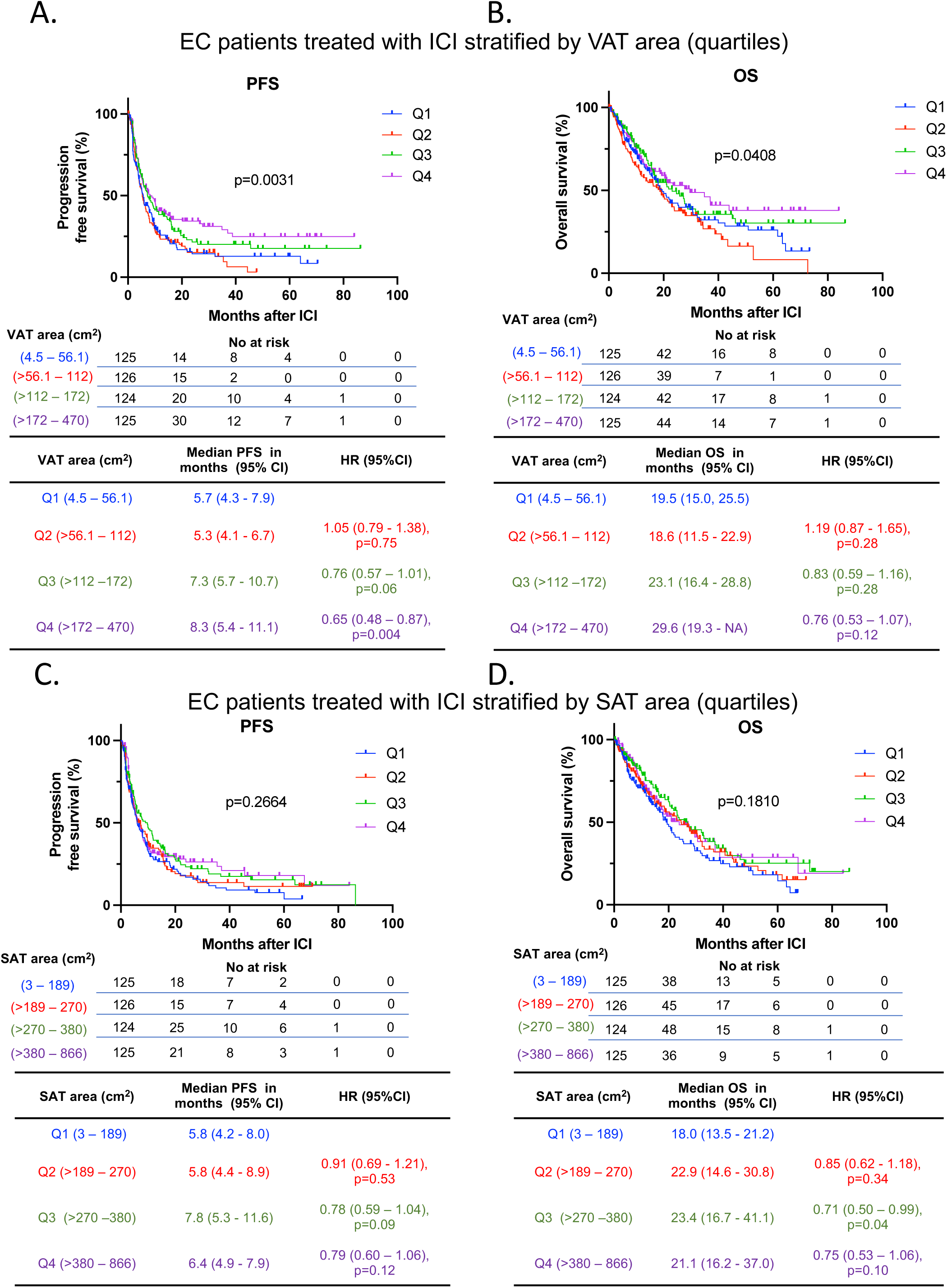
Survival outcomes after ICI in EC stratified by VAT and SAT area quartiles. Kaplan-Meier curves for (A) PFS and (B) OS in patients with EC following ICI treatment stratified by VAT area quartiles (Q1, 4.5 – 56.1 cm^2^ in blue; Q2, >56.1 – 112 cm^2^ in red; Q3, >112 – 172 cm^2^ in green; Q4, >172 – 470 cm^2^ in purple) (n=500). Kaplan-Meier curves for (C) PFS and (D) OS in patients with EC following ICI treatment stratified by SAT area quartiles SAT area quartiles (Q1, 3 – 189 cm^2^ in blue; Q2, >189 – 270 cm^2^ in red; Q3, >270 – 380 cm^2^ in green; Q4, >380 – 866 cm^2^ in purple) (n=500). The P values in the PFS and OS plots were calculated using a log-rank test. HRs and 95% CIs were calculated using Q1 as a reference. BMI, body mass index; OS, overall survival; PFS, progression free survival; EC, endometrial cancer; ICI, immune checkpoint inhibitor; VAT, visceral adipose tissue; SAT, subcutaneous adipose tissue; HR, Hazard ratio; CI, confidence interval.

**Supplemental Figure 3.**
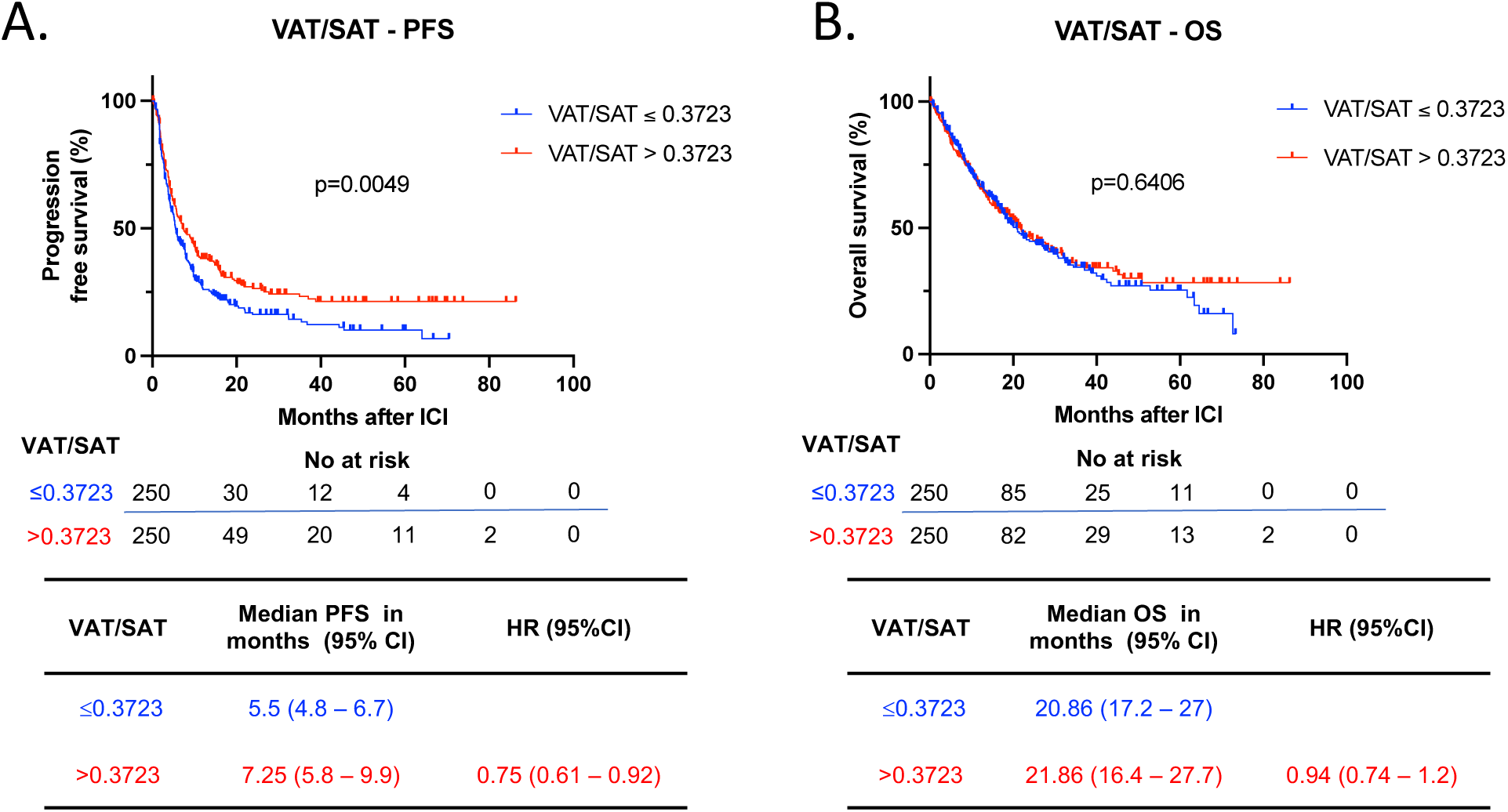
Survival outcomes after ICI in EC stratified by VAT/SAT ratio. Kaplan-Meier curves for (A) PFS and (B) OS in patients with EC following ICI treatment stratified by VAT/SAT ratio (n=500) (Low VAT/SAT ratio: ≤0.3723 in blue; High VAT/SAT ratio: >0.3723 in red). Patients were categorized as low or high VAT /SAT based on the median SAT and VAT of the entire cohort. The P values in the in PFS and OS plots were calculated using a log-rank test. BMI, body mass index; OS, overall survival; PFS, progression free survival; EC, endometrial cancer; ICI, immune checkpoint inhibitor; VAT, visceral adipose tissue; SAT, subcutaneous adipose tissue; HR, Hazard ratio; CI, confidence interval.

**Supplemental Figure 4.**
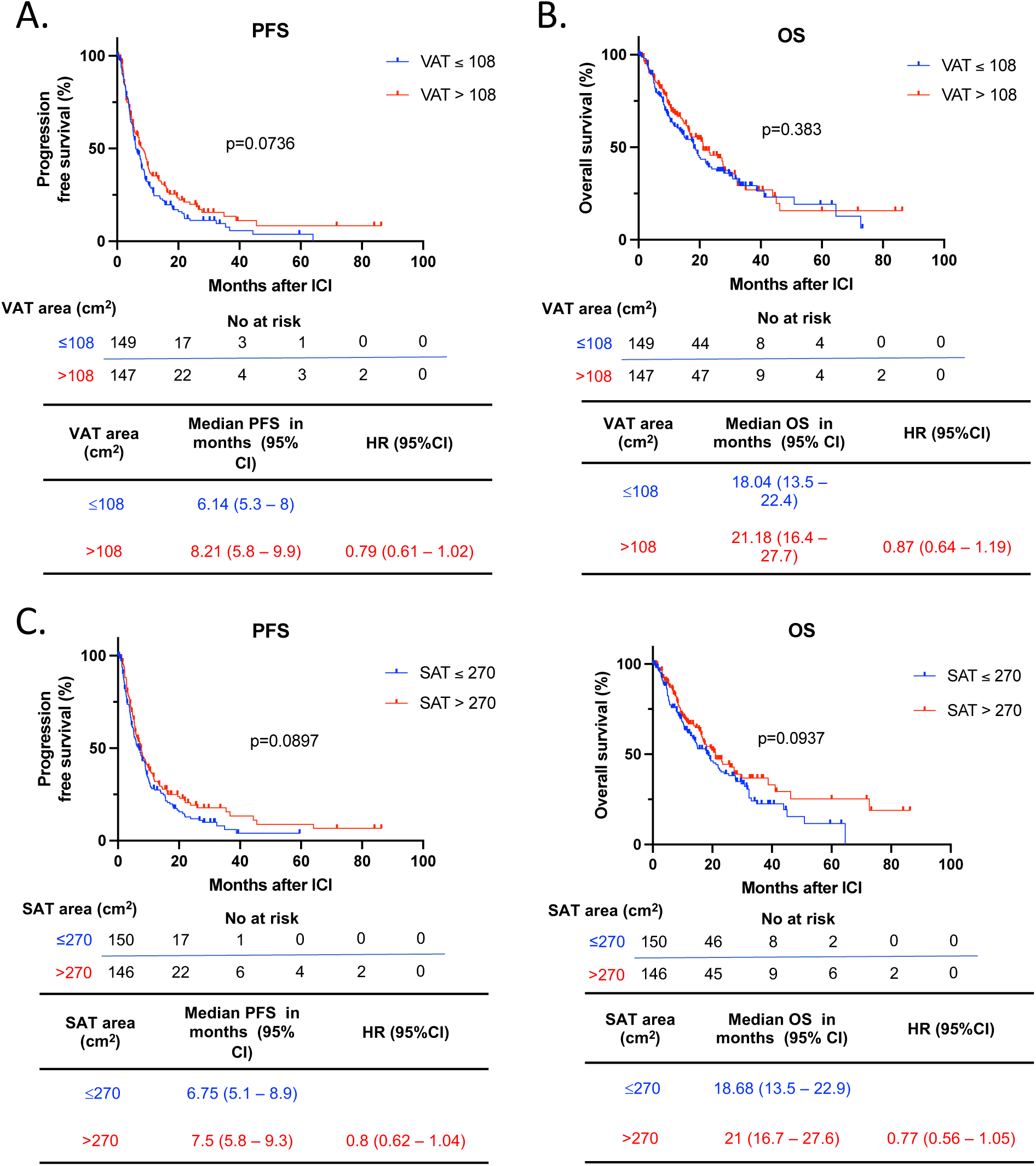
Survival outcomes after Pembrolizumab and Lenvatinib in EC stratified by VAT and SAT area. Kaplan-Meier curves for (A) PFS and (B) OS in patients with EC following ICI treatment stratified by low and high VAT area (n=296) (Low VAT area: ≤108 cm^2^ in blue; High VAT area: >108 cm^2^ in red). Kaplan-Meier curves for (C) PFS and (D) OS in patients with EC following ICI treatment stratified by low and high SAT area (Low SAT area: ≤270 cm^2^ in blue; High SAT area: >270 cm^2^ in red) (n=500). Patients were categorized as low or high VAT /SAT based on the median SAT and VAT of the patient sub-group treated with Pembrolizumab + Lenvatinib. The P values in the PFS and OS plots were calculated using a log-rank test. BMI, body mass index; OS, overall survival; PFS, progression free survival; EC, endometrial cancer; ICI, immune checkpoint inhibitor; VAT, visceral adipose tissue; SAT, subcutaneous adipose tissue; HR, Hazard ratio; CI, confidence interval.

**Supplemental Figure 5.**
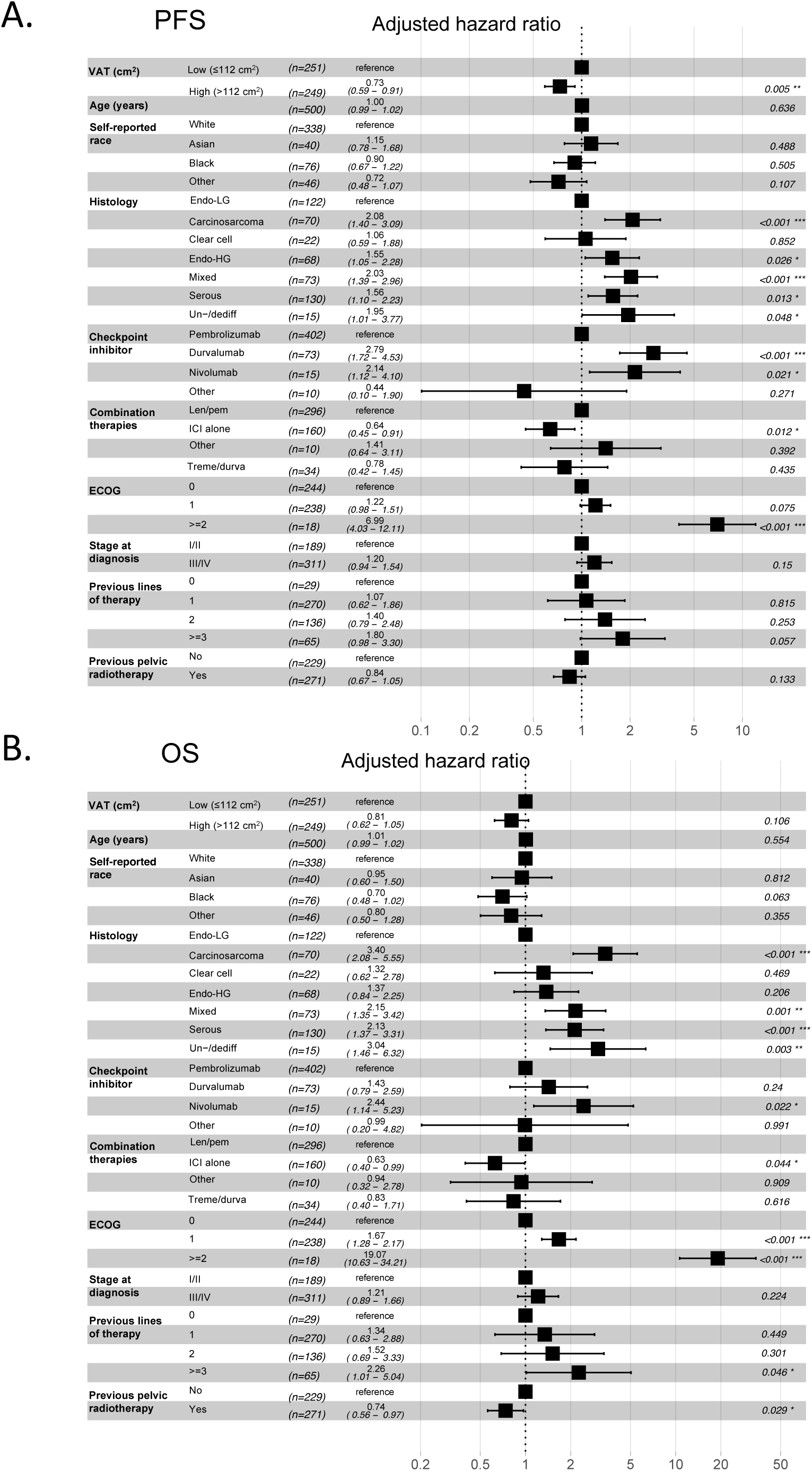
Multivariable Cox regression analysis of VAT and other clinical variables associated with responses to ICI in EC patients. Forest plots of adjusted HRs and 95% CIs for patients with low VAT (reference group) compared to high VAT for (A) PFS and (B) OS (n=500). Analysis was adjusted for age, self-reported race, histology, type of checkpoint inhibitor, combination therapies, baseline performance status, stage at diagnosis, prior lines of therapy, and previous pelvic radiation. BMI, body mass index; OS, overall survival; PFS, progression free survival; EC, endometrial cancer; ICI, immune checkpoint inhibitor; VAT, visceral adipose tissue; Endo-LG, endometrial low grade; Endo-HG, endometrial high grade; Un-/dediff, Un-/dedifferentiated; Len/pem, Lenvatinib/pembrolizumab; Treme/durva, Tremelimumab/durvalumab; HR, Hazard ratio; CI, confidence interval.

**Supplemental Figure 6.**
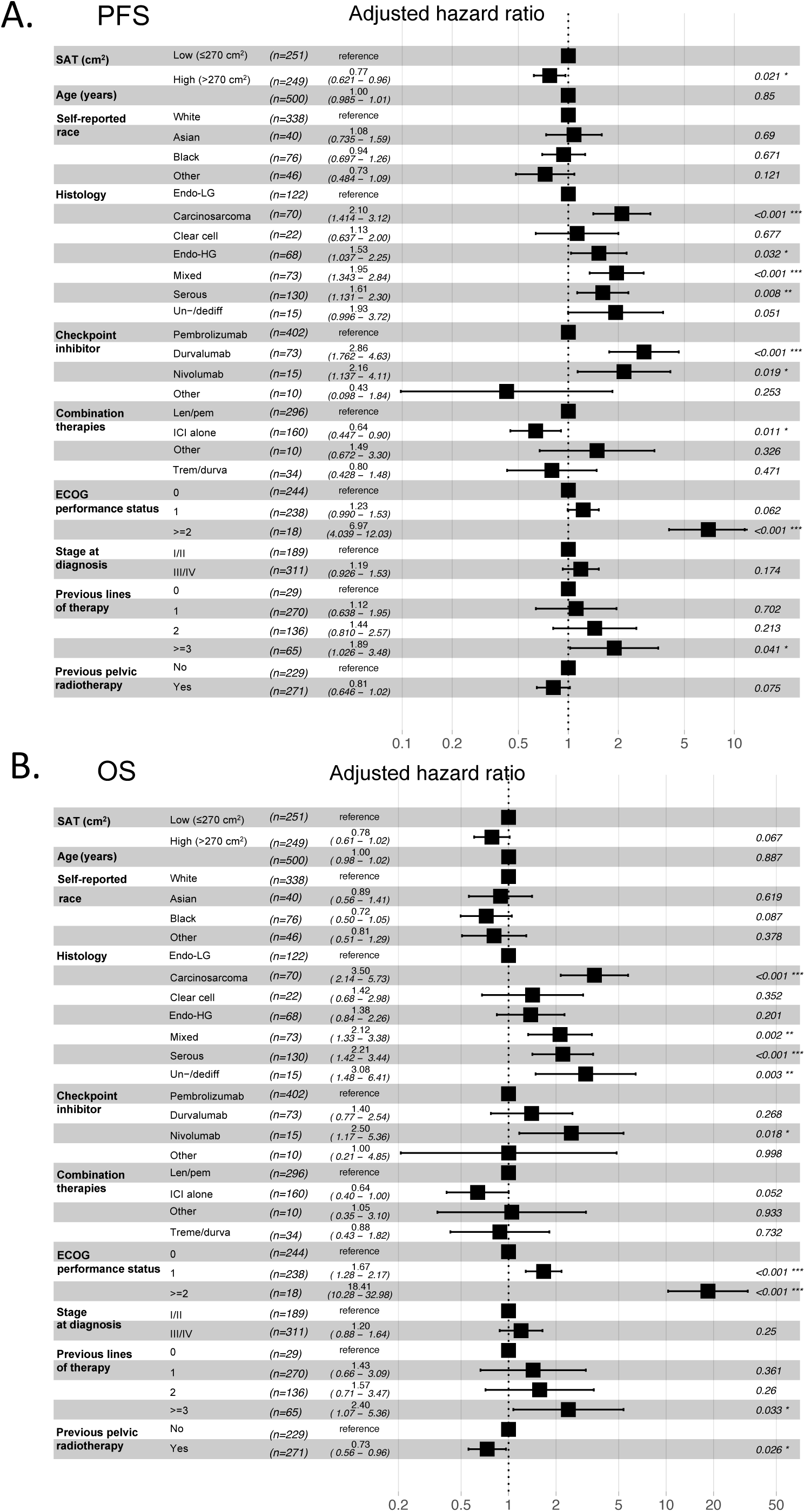
Multivariable Cox regression analysis of SAT and other clinical variables associated with responses to ICI in EC patients. Forest plots of adjusted HRs and 95% Cis for patients with low SAT (reference group) compared to high SAT for (A) PFS and (B) OS (n=500). Analysis was adjusted for age, self-reported race, histology, type of checkpoint inhibitor, combination therapies, baseline performance status, stage at diagnosis, prior lines of therapy, and previous pelvic radiation. BMI, body mass index; OS, overall survival; PFS, progression free survival; EC, endometrial cancer; ICI, immune checkpoint inhibitor; SAT, subcutaneous adipose tissue; Endo-LG, endometrial low grade; Endo-HG, endometrial high grade; Un-/dediff, Un-/dedifferentiated; Len/pem, Lenvatinib/pembrolizumab; Treme/durva, Tremelimumab/durvalumab; HR, Hazard ratio; CI, confidence interval.

**Supplemental Figure 7.**
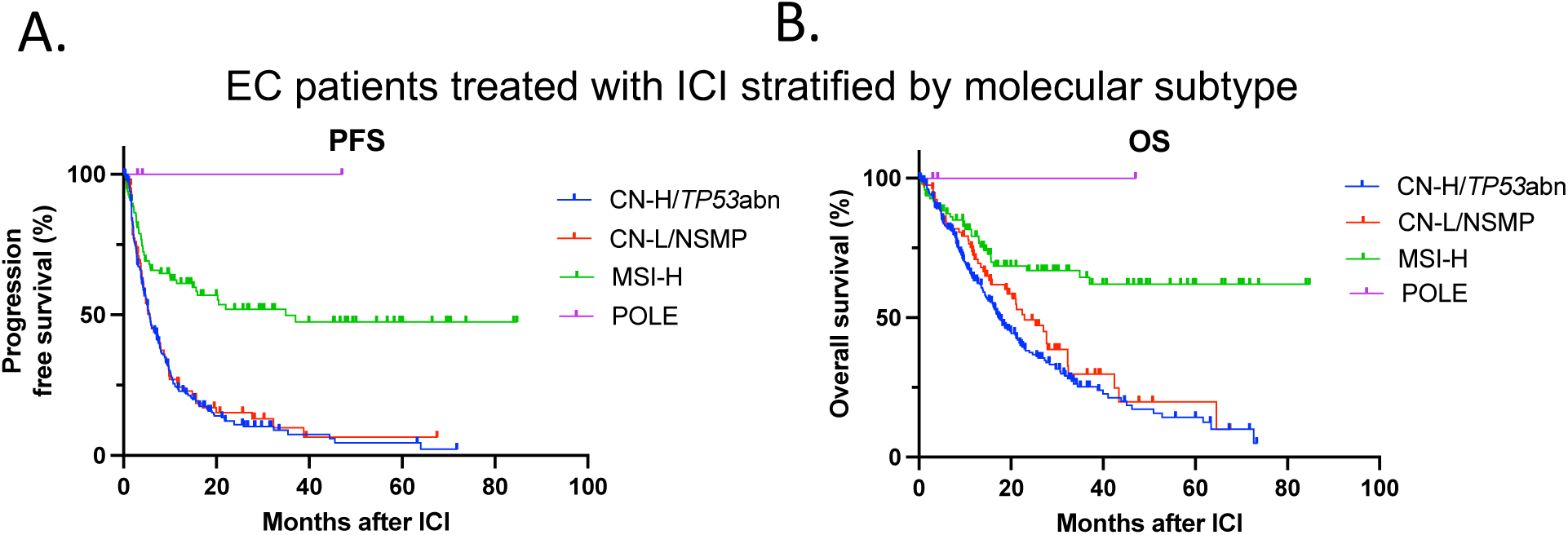
Survival outcomes in patients with EC stratified by molecular classification after ICI treatment. Kaplan-Meier curves for (A) OS and (B) PFS in patients with EC following ICI treatment stratified by molecular subtype (n=437). OS, overall survival; PFS, progression free survival; EC, endometrial cancer; ICI, immune checkpoint inhibitor; CN-H/*TP53*abn, copy number-high/*TP53* abnormal; CN-L/NSMP, copy number-low/no specific molecular profile; MSI-H, microsatellite instability-high.

**Supplemental Figure 8.**
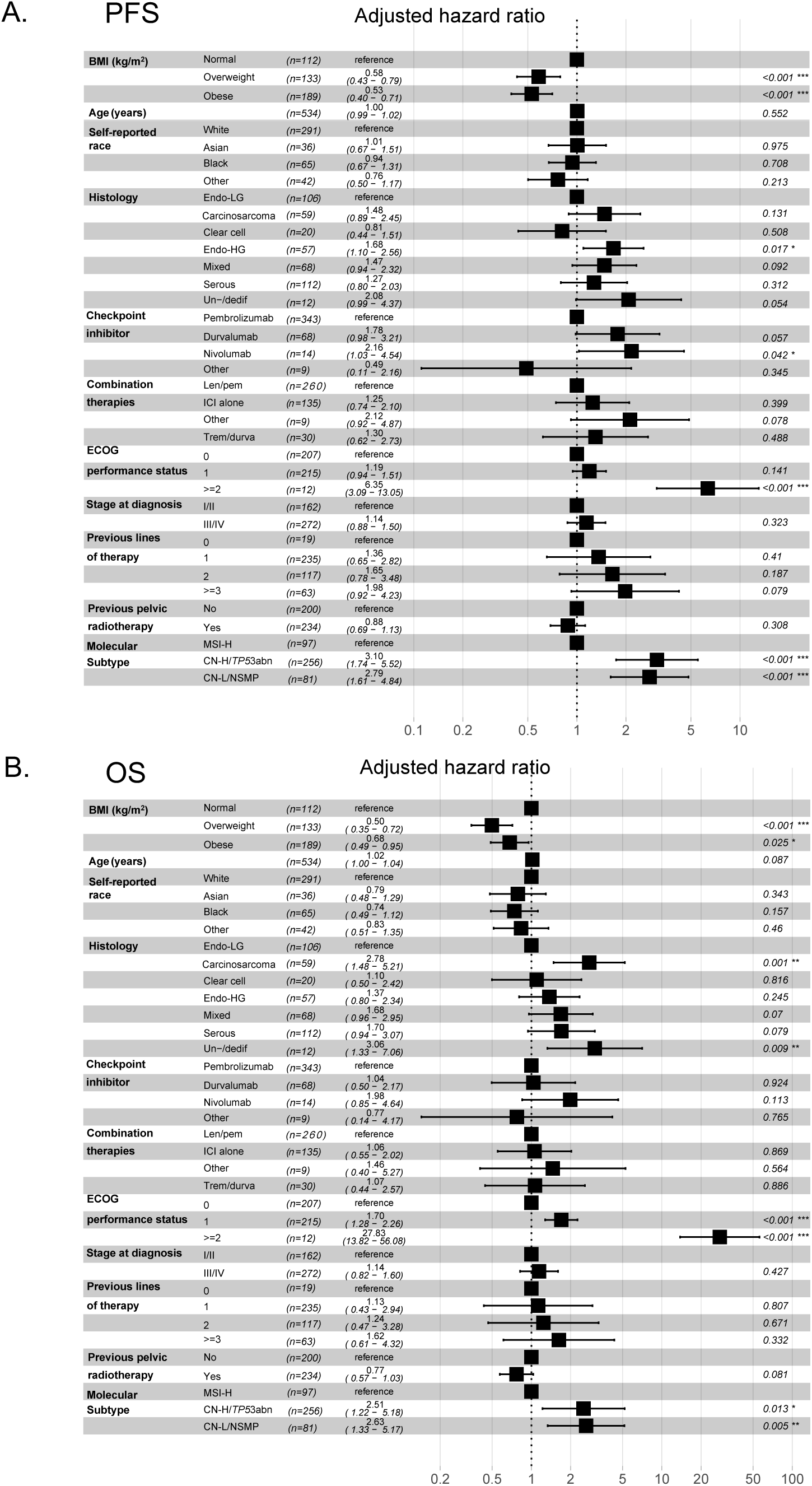
Multivariable regression analysis of clinical and molecular signatures associated with responses to ICI in EC patients. Forest plots of adjusted HRs and 95% CIs for patients with normal BMI (18.5 – 24.9 kg/m^2^) (reference group) compared to overweight (BMI 25 – 29.9 kg/m^2^) and obese (BMI > 30 kg/m^2^) patients for (A) PFS and (B) (OS) (n=434). There were a limited number of POLE patients (n=3), so they were not included in the analysis. Analysis was adjusted for age, self-reported race, histology, type of checkpoint inhibitor, combination therapies, baseline performance status, stage at diagnosis, prior lines of therapy, previous pelvic radiation, and molecular subtype. BMI, body mass index; OS, overall survival; PFS, progression free survival; EC, endometrial cancer; ICI, immune checkpoint inhibitor; Endo-LG, endometrial low grade; Endo-HG, endometrial high grade; Un-/dediff, Un-/dedifferentiated; Len/pem, Lenvatinib/pembrolizumab; Treme/durva, Tremelimumab/durvalumab; CN-H/*TP53*abn, copy number-high/*TP53* abnormal; CN-L/NSMP, copy number-low/no specific molecular profile; MSI-H, microsatellite instability-high; HR, Hazard ratio; CI, confidence interval.

**Supplemental Figure 9.**
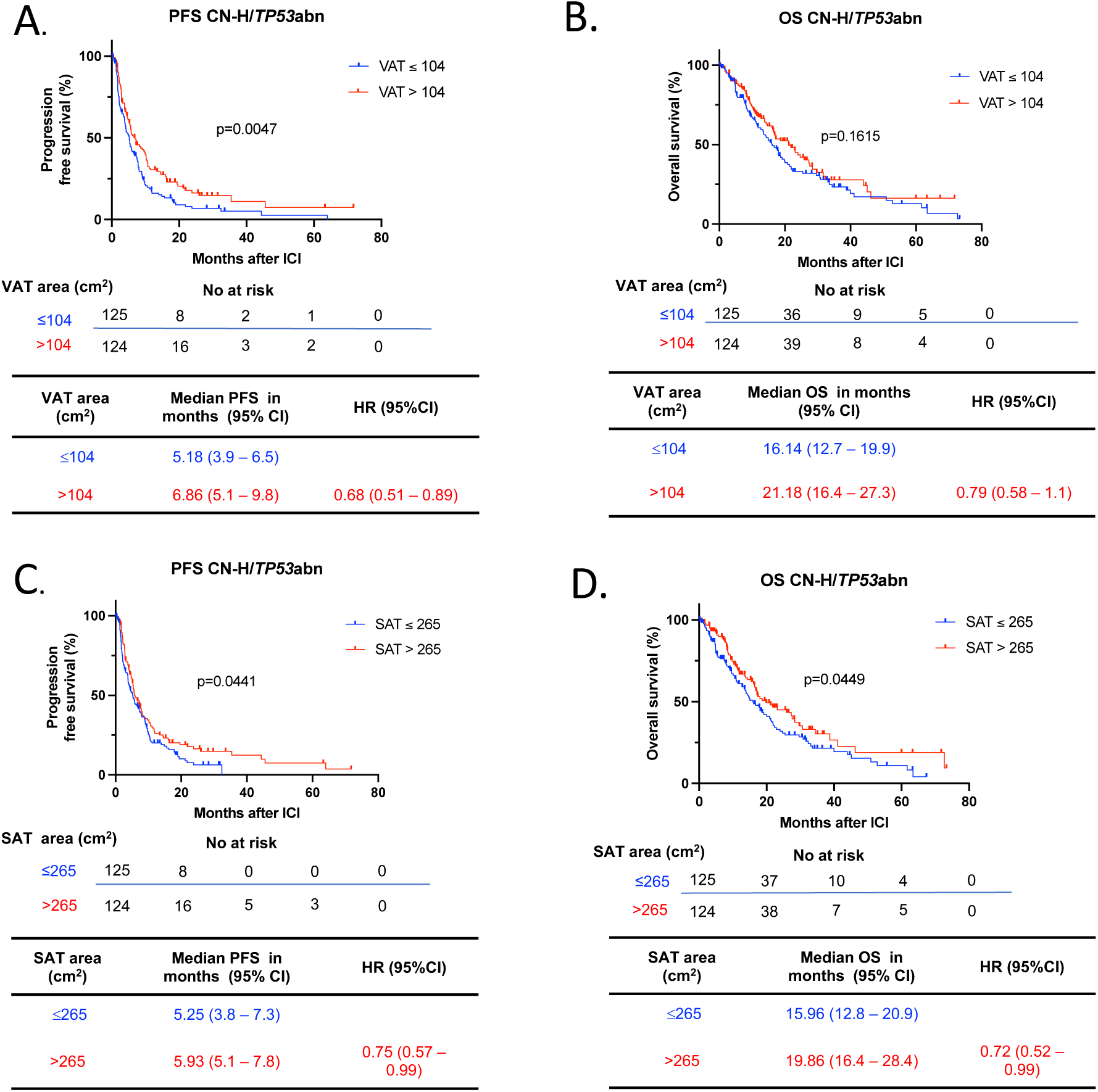
Survival outcomes in CN-H/*TP53*abnl EC stratified by VAT and SAT area. Kaplan-Meier curves for (A) PFS and (B) OS in patients with EC following ICI treatment stratified by low and high VAT area (n=249) (Low VAT area: ≤104 cm2 in blue; High VAT area: >104 cm2 in red). Kaplan-Meier curves for (C) PFS and (D) OS in patients with EC following ICI treatment stratified by low and high SAT area (Low SAT area: ≤265 cm2 in blue; High SAT area: >265 cm2 in red) (n=249). Patients were categorized as low or high VAT /SAT based on the median SAT and VAT area of the CN-H/TP53abn EC patients with available radiological data. The P values were calculated using a log-rank test. HRs and 95% CIs for overweight and obese patients were calculated using normal weight as a reference. BMI, body mass index; VAT, visceral adipose tissue; SAT, subcutaneous adipose tissue; OS, overall survival; PFS, progression free survival; EC, endometrial cancer; ICI, immune checkpoint inhibitor; CN-H/TP53abnl, copy number-high/TP53abnormal; HR, Hazard ratio; CI, confidence interval.

**Supplemental Figure 10.**
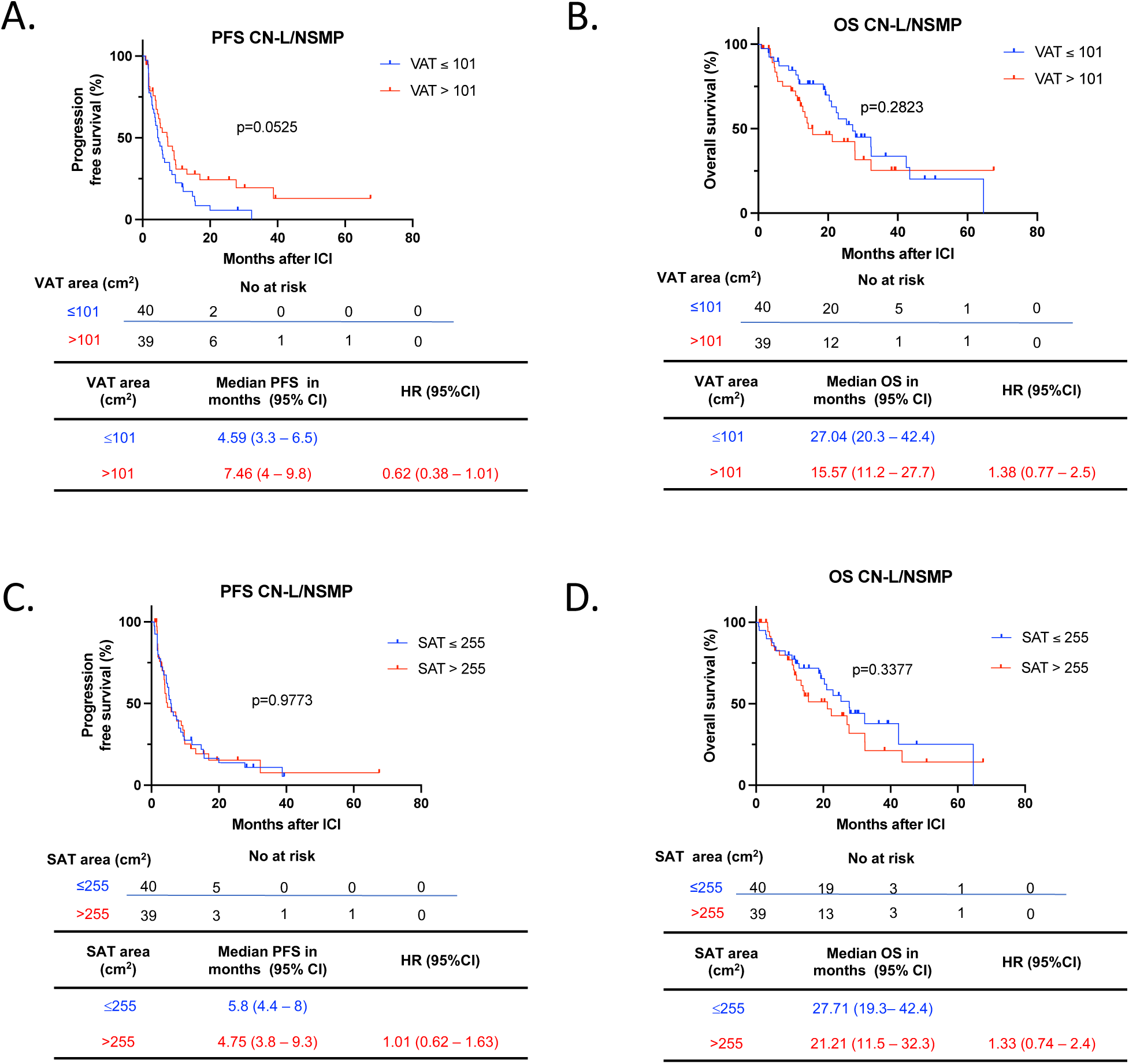
Survival outcomes in CN-L/NSMP EC stratified by VAT and SAT area. Kaplan-Meier curves for (A) PFS and (B) OS in patients with EC following ICI treatment stratified by low and high VAT area (n=79) (Low VAT area: ≤101 cm2 in blue; High VAT area: >101 cm2 in red). Kaplan-Meier curves for (C) PFS and (D) OS in patients with EC following ICI treatment stratified by low and high SAT area (Low SAT area: ≤255 cm2 in blue; High SAT area: >255 cm2 in red) (n=79). Patients were categorized as low or high VAT /SAT based on the median SAT and VAT area of the CN-L/NSMP EC patients with available radiological data. The P values were calculated using a log-rank test. HRs and 95% CIs for overweight and obese patients were calculated using normal weight as a reference. BMI, body mass index; VAT, visceral adipose tissue; SAT, subcutaneous adipose tissue; OS, overall survival; PFS, progression free survival; EC, endometrial cancer; ICI, immune checkpoint inhibitor; CN-L/NSMP, copy number-low/no specific molecular profile; HR, Hazard ratio; CI, confidence interval.

**Supplemental Figure 11.**
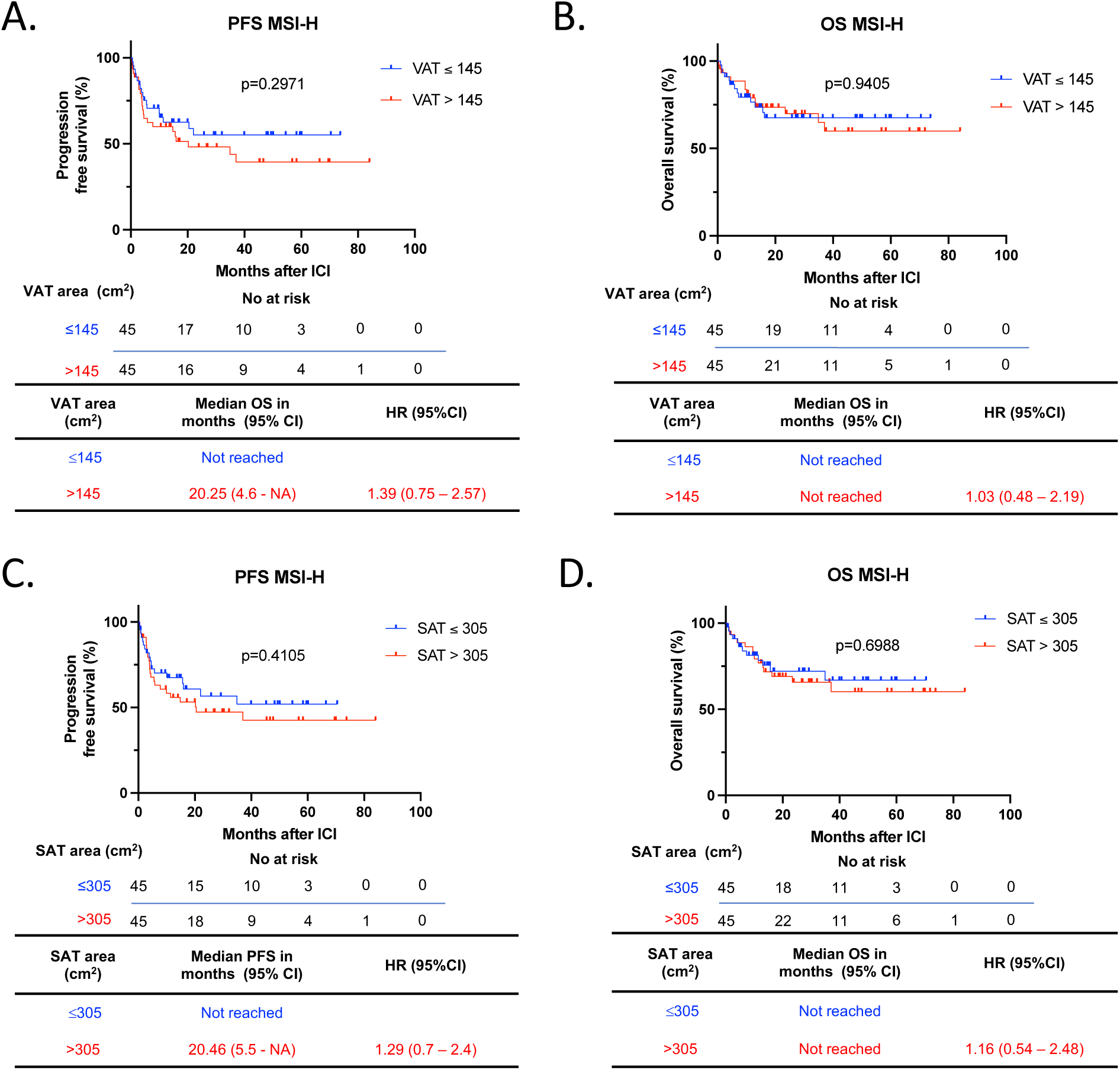
Survival outcomes in MSI-H EC stratified by VAT and SAT area. Kaplan-Meier curves for (A) PFS and (B) OS in patients with EC following ICI treatment stratified by low and high VAT area (n=90) (Low VAT area: ≤145 cm2 in blue; High VAT area: >145 cm2 in red). Kaplan-Meier curves for (C) PFS and (D) OS in patients with EC following ICI treatment stratified by low and high SAT area (Low SAT area: ≤305 cm2 in blue; High SAT area: >305 cm2 in red) (n=90). Patients were categorized as low or high VAT /SAT based on the median SAT and VAT area of the MSI-H EC patients with available radiological data. The P values were calculated using a log-rank test. HRs and 95% CIs for overweight and obese patients were calculated using normal weight as a reference. BMI, body mass index; VAT, visceral adipose tissue; SAT, subcutaneous adipose tissue; OS, overall survival; PFS, progression free survival; EC, endometrial cancer; ICI, immune checkpoint inhibitor; MSI-H, microsatellite instability-high; HR, Hazard ratio; CI, confidence interval.

**Supplemental Figure 12.**
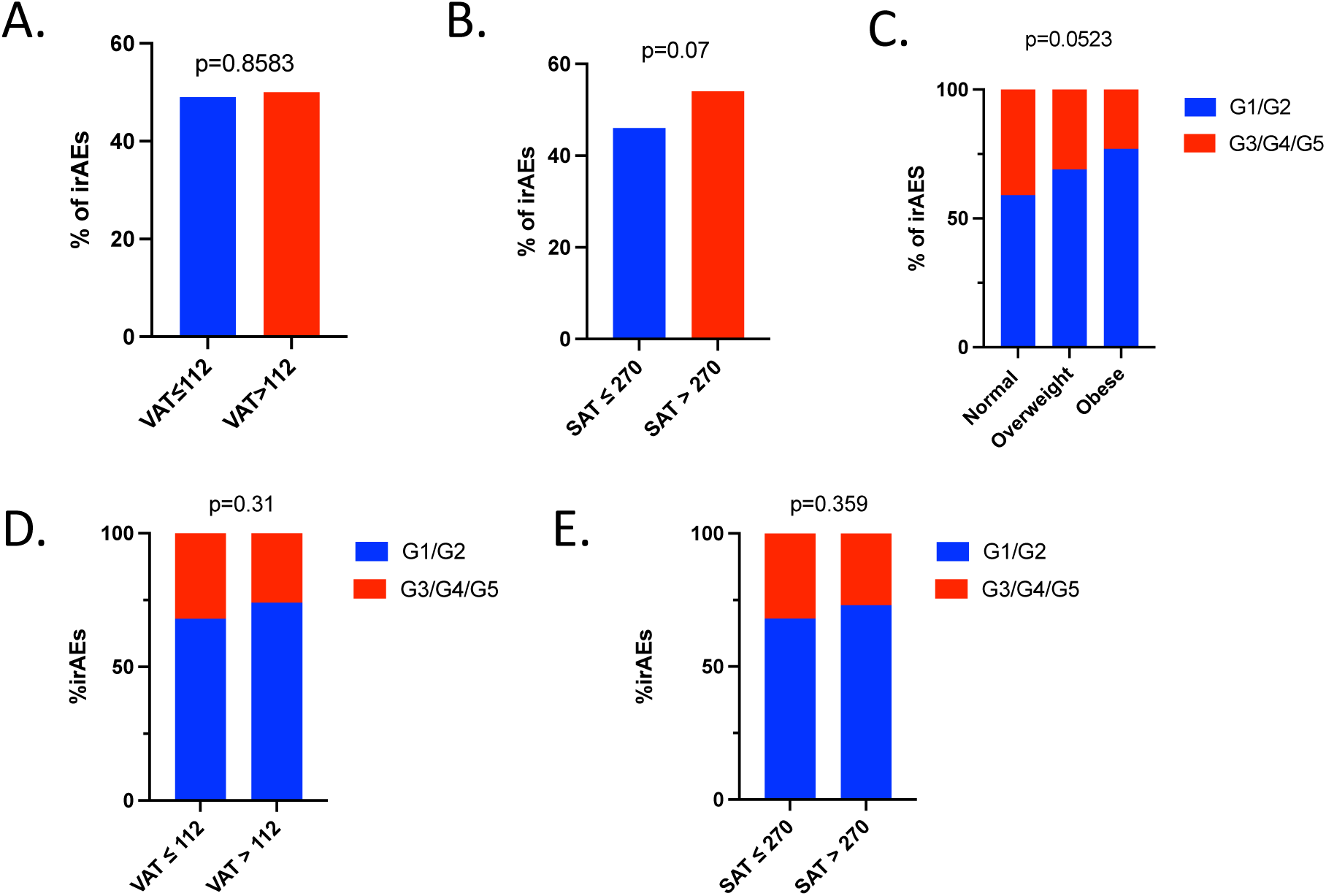
Incidence of irAEs in EC patients after treatment with ICI stratified by BMI, VAT and SAT. Percentage of irAEs stratified by (A) VAT area and (B) SAT area (n=500). Percentage of mild/moderate (G1/G2) irAEs vs severe (G3/G4) irAEs stratified by (C) BMI (D) VAT area, and (E) SAT area (n=500). The P value in the bar graph was calculated using chi-squared test. BMI, body mass index; VAT, visceral adipose tissue; SAT, subcutaneous adipose tissue; EC, endometrial cancer; ICI, immune checkpoint inhibitor; iRAEs, immune related adverse events; G1/G2, Grade 1 and Grade 2; G3/G4/G5, Grade 3, Grade 4 and Grade 5.

**Supplemental Table 1.** Clinical characteristics of EC patients with available molecular subtype. BMI, body mass index; ECOG, Eastern Cooperative Oncology Group; CPN-H/*TP53*abn, copy number-high/*TP53*abnormal; CN-L/NSMP, copy number-low/no specific molecular profile; MSI-H, microsatellite instability high; VAT, visceral adipose tissue; SAT, subcutaneous adipose tissue. P values in table come from Kruskal-Wallis, chi-squared or Fisher’s exact tests.

|  | All n=437 | Normal BMI<br>n=114 | Overweight<br>n=133 | Obese<br>n=190 | p value |
| --- | --- | --- | --- | --- | --- |
| **Median age (range)** | 67 (40 – 94) | 67 (41 – 89) | 68 (43 – 94) | 66 (40 – 88) | 0.02 |
| **Median BMI, kg/m<sup>2</sup> (range)** | 28.8 (18.5 – 59.4) | 22.5 (18.5 – 24.9) | 27 (25 – 29.9) | 35.1 (30 – 59.4) | < 0.0001 |
| **Self-reported race - No (%)** |  |  |  |  |  |
| • White: | 293 (67) | 75 (66) | 99 (74) | 119 (63) | 0.01 |
| • Black: | 65 (15) | 13 (11) | 17 (13) | 35 (18) |  |
| • Asian: | 36 (8) | 14 (12) | 12 (9) | 10 (5) |  |
| • Other: | 43 (10) | 12 (11) | 5 (4) | 26 (14) |  |
| **Histology – No (%)** |  |  |  |  |  |
| • Endometroid | 166 (38) | 38 (33) | 48 (36) | 80 (42) | 0.12 |
| ○ Low grade (1,2) | 107 (24) | 27 (24) | 33 (25) | 47 (25) |  |
| ○ High grade (3) | 59 (14) | 11 (10) | 15 (11) | 33 (17) |  |
| • Serous | 112 (26) | 23 (20) | 40 (30) | 49 (26) |  |
| • Mixed/high-grade NOS | 68 (16) | 17 (15) | 20 (15) | 31 (16) |  |
| • Carcinosarcoma | 59 (14) | 22 (19) | 17 (13) | 20 (11) |  |
| • Clear Cell | 20 (5) | 7 (6) | 6 (5) | 7 (4) |  |
| • Un- / Dedifferentiated | 12 (3) | 7 (6) | 2 (2) | 3 (2) |  |
| **Checkpoint inhibitor – No (%)** |  |  |  |  |  |
| • Pembrolizumab: | 346 (79) | 83 (73) | 106 (80) | 157 (83) | 0.34 |
| • Durvalumab: | 68 (16) | 21 (18) | 22 (17) | 25 (13) |  |
| • Nivolumab: | 14 (3) | 6 (5) | 4 (3) | 4 (2) |  |
| • Other: | 9 (2) | 4 (4) | 1 (1) | 4 (2) |  |
| **Combination therapies – No (%)** |  |  |  |  |  |
| • Lenvatinib/pembrolizumab: | 260 (59) | 62 (54) | 80 (60) | 118 (62) | 0.76 |
| • Tremelimumab/durvalumab: | 30 (7) | 10 (9) | 8 (6) | 12 (6) |  |
| • ICI alone: | 138 (32) | 38 (33) | 43 (32) | 57 (30) |  |
| • Other combination: | 9 (2) | 4 (4) | 2 (2) | 3 (2) |  |
| **ECOG Performance Status– No (%)** |  |  |  |  |  |
| • 0: | 208 (48) | 56 (49) | 73 (55) | 79 (42) | 0.18 |
| • 1: | 217 (50) | 55 (48) | 56 (42) | 106 (56) |  |
| • 2-3 | 12 (3) | 3 (3) | 4 (3) | 5 (3) |  |
| **Stage at diagnosis (1,2 vs 3,4) – No (%)** |  |  |  |  |  |
| • 1,2: | 165 (38) | 41(36) | 50 (38) | 74 (39) | 0.87 |
| • 3,4: | 272 (62) | 73 (64) | 83 (62) | 116 (61) |  |
| **Previous lines of therapy – No (%)** |  |  |  |  |  |
| • 0 | 20 (5) | 8 (7) | 7 (5) | 5 (3) | 0.17 |
| • 1 | 236 (54) | 50 (44) | 73 (55) | 113 (59) |  |
| • 2 | 118 (27) | 35 (31) | 36 (27) | 47 (25) |  |
| • ≥ 3 | 63 (14) | 21 (18) | 17 (13) | 25 (13) |  |
| **Previous pelvic radiotherapy – No (%)** |  |  |  |  |  |
| • Yes | 237 (54) | 57 (50) | 80 (60) | 100 (53) | 0.24 |
| • No | 200 (46) | 57 (50) | 53 (40) | 90 (47) |  |
| **Molecular subtype – No (%)** |  |  |  |  |  |
| • CN-H/*TP53*abn | 256 (59) | 66 (58) | 77 (58) | 113 (59) | 0.06 |
| • MSI-H | 97 (22) | 21 (18) | 25 (19) | 51 (27) |  |
| • CN-L/NSMP | 81 (19) | 25 (22) | 31 (23) | 25 (13) |  |
| • POLE | 3 (0.7) | 2 (2) | 0 (0) | 1 (0.5) |  |

**Supplemental Table 2.** irAEs per organ system in EC patients after treatment with ICI stratified by BMI. Absolute number and percentage of irAEs per organ system across BMI categories Normal - BMI: 18.5 – 24.9 kg/m^2^; Overweight – BMI 25 – 29.9 kg/m^2^; Obese – BMI > 30 kg/m^2^. P-values in were calculated with Fisher’s exact test. BMI, body mass index; EC, endometrial cancer; ICI, immune checkpoint inhibitor; irAEs, immune related adverse events.

| BMI (Kg/m<sup>2</sup>) | Pneumonitis | Myocarditis | Neurological | Ocular | Hematologic |
| --- | --- | --- | --- | --- | --- |
| **Normal**<br>**(18.5 – 24.9) – No (%)** | 1 (0.8) | 0 (0) | 3 (2) | 0 (0) | 1 (0.8) |
| **Overweight**<br>**(25 – 29.9) – No (%)** | 1 (0.6) | 2 (1) | 0 (0) | 1 (0.6) | 1 (0.6) |
| **Obese**<br>**(≥ 30) – No (%)** | 2 (0.9) | 1 (0.4) | 2 (0.9) | 1 (0.4) | 0 (0) |
| **Total - No (%)** | 4 (0.7) | 3 (0.6) | 5 (1) | 2 (0.4) | 2 (0.4) |
| **p value** | 1 | 0.5 | 0.1 | 1 | 0.3 |

## REFERENCES

1. Somasegar S, Bashi A, Lang SM, Liao CI, Johnson C, Darcy KM, et al. Trends in Uterine Cancer Mortality in the United States: A 50-Year Population-Based Analysis. Obstet Gynecol. 2023;142(4):978–86.

2. Global BMIMC, Di Angelantonio E, Bhupathiraju Sh N, Wormser D, Gao P, Kaptoge S, et al. Body-mass index and all-cause mortality: individual-participant-data meta-analysis of 239 prospective studies in four continents. Lancet. 2016;388(10046):776–86.

3. Makker V, MacKay H, Ray-Coquard I, Levine DA, Westin SN, Aoki D, et al. Endometrial cancer. Nat Rev Dis Primers. 2021;7(1):88.

4. Lauby-Secretan B, Scoccianti C, Loomis D, Grosse Y, Bianchini F, Straif K, et al. Body Fatness and Cancer--Viewpoint of the IARC Working Group. N Engl J Med. 2016;375(8):794–8.

5. Secord AA, Hasselblad V, Von Gruenigen VE, Gehrig PA, Modesitt SC, Bae-Jump V, et al. Body mass index and mortality in endometrial cancer: A systematic review and meta-analysis. Gynecol Oncol. 2016;140(1):184–90.

6. Crosbie EJ, Kitson SJ, McAlpine JN, Mukhopadhyay A, Powell ME, and Singh N. Endometrial cancer. Lancet. 2022;399(10333):1412–28.

7. Hotamisligil GS. Inflammation and metabolic disorders. Nature. 2006;444(7121):860-7.

8. Bouwman F, Smits A, Lopes A, Das N, Pollard A, Massuger L, et al. The impact of BMI on surgical complications and outcomes in endometrial cancer surgery--an institutional study and systematic review of the literature. Gynecol Oncol. 2015;139(2):369–76.

9. Hamoud BH, Sima RM, Vacaroiu IA, Georgescu MT, Bobirca A, Gaube A, et al. The Evolving Landscape of Immunotherapy in Uterine Cancer: A Comprehensive Review. Life (Basel). 2023;13(7).

10. Mirza MR, Chase DM, Slomovitz BM, dePont Christensen R, Novak Z, Black D, et al. Dostarlimab for Primary Advanced or Recurrent Endometrial Cancer. N Engl J Med. 2023;388(23):2145–58.

11. Eskander RN, Sill MW, Beffa L, Moore RG, Hope JM, Musa FB, et al. Pembrolizumab plus Chemotherapy in Advanced Endometrial Cancer. N Engl J Med. 2023;388(23):2159–70.

12. Makker V, Colombo N, Casado Herraez A, Santin AD, Colomba E, Miller DS, et al. Lenvatinib plus Pembrolizumab for Advanced Endometrial Cancer. N Engl J Med. 2022;386(5):437–48.

13. O’Malley DM, Bariani GM, Cassier PA, Marabelle A, Hansen AR, De Jesus Acosta A, et al. Pembrolizumab in Patients With Microsatellite Instability-High Advanced Endometrial Cancer: Results From the KEYNOTE-158 Study. J Clin Oncol. 2022;40(7):752–61.

14. Lysaght J. The ‘obesity paradox’ in action with cancer immunotherapy. Nat Rev Endocrinol. 2019;15(3):132–3.

15. McQuade JL, Hammers H, Furberg H, Engert A, Andre T, Blumenschein G, Jr., et al. Association of Body Mass Index With the Safety Profile of Nivolumab With or Without Ipilimumab. JAMA Oncol. 2023;9(1):102–11.

16. McQuade JL, Daniel CR, Hess KR, Mak C, Wang DY, Rai RR, et al. Association of body-mass index and outcomes in patients with metastatic melanoma treated with targeted therapy, immunotherapy, or chemotherapy: a retrospective, multicohort analysis. Lancet Oncol. 2018;19(3):310–22.

17. Lalani AA, Bakouny Z, Farah S, Donskov F, Dudani S, Heng DYC, et al. Assessment of Immune Checkpoint Inhibitors and Genomic Alterations by Body Mass Index in Advanced Renal Cell Carcinoma. JAMA Oncol. 2021;7(5):773–5.

18. Ged Y, Sanchez A, Patil S, Knezevic A, Stein E, Petruzella S, et al. Associations between Pretreatment Body Composition Features and Clinical Outcomes among Patients with Metastatic Clear Cell Renal Cell Carcinoma Treated with Immune Checkpoint Blockade. Clin Cancer Res. 2022;28(23):5180–9.

19. Yoo SK, Chowell D, Valero C, Morris LGT, and Chan TA. Outcomes Among Patients With or Without Obesity and With Cancer Following Treatment With Immune Checkpoint Blockade. JAMA Netw Open. 2022;5(2):e220448.

20. Kichenadasse G, Miners JO, Mangoni AA, Rowland A, Hopkins AM, and Sorich MJ. Association Between Body Mass Index and Overall Survival With Immune Checkpoint Inhibitor Therapy for Advanced Non-Small Cell Lung Cancer. JAMA Oncol. 2020;6(4):512–8.

21. Antoun S, Lanoy E, Ammari S, Farhane S, Martin L, Robert C, et al. Protective effect of obesity on survival in cancers treated with immunotherapy vanishes when controlling for type of cancer, weight loss and reduced skeletal muscle. Eur J Cancer. 2023;178:49–59.

22. Gonzalez MC, Correia M, and Heymsfield SB. A requiem for BMI in the clinical setting. Curr Opin Clin Nutr Metab Care. 2017;20(5):314–21.

23. Bradshaw PT. Body composition and cancer survival: a narrative review. Br J Cancer. 2023.

24. Irlbeck T, Massaro JM, Bamberg F, O’Donnell CJ, Hoffmann U, and Fox CS. Association between single-slice measurements of visceral and abdominal subcutaneous adipose tissue with volumetric measurements: the Framingham Heart Study. Int J Obes (Lond). 2010;34(4):781–7.

25. Kaess BM, Pedley A, Massaro JM, Murabito J, Hoffmann U, and Fox CS. The ratio of visceral to subcutaneous fat, a metric of body fat distribution, is a unique correlate of cardiometabolic risk. Diabetologia. 2012;55(10):2622–30.

26. Britton KA, Massaro JM, Murabito JM, Kreger BE, Hoffmann U, and Fox CS. Body fat distribution, incident cardiovascular disease, cancer, and all-cause mortality. J Am Coll Cardiol. 2013;62(10):921–5.

27. Rios-Doria E, Momeni-Boroujeni A, Friedman CF, Selenica P, Zhou Q, Wu M, et al. Integration of clinical sequencing and immunohistochemistry for the molecular classification of endometrial carcinoma. Gynecol Oncol. 2023;174:262–72.

28. Cancer Genome Atlas Research N, Kandoth C, Schultz N, Cherniack AD, Akbani R, Liu Y, et al. Integrated genomic characterization of endometrial carcinoma. Nature. 2013;497(7447):67–73.

29. Conroy M, and Naidoo J. Immune-related adverse events and the balancing act of immunotherapy. Nat Commun. 2022;13(1):392.

30. Osorio JC, Ni A, Chaft JE, Pollina R, Kasler MK, Stephens D, et al. Antibody-mediated thyroid dysfunction during T-cell checkpoint blockade in patients with non-small-cell lung cancer. Ann Oncol. 2017;28(3):583–9.

31. Maher VE, Fernandes LL, Weinstock C, Tang S, Agarwal S, Brave M, et al. Analysis of the Association Between Adverse Events and Outcome in Patients Receiving a Programmed Death Protein 1 or Programmed Death Ligand 1 Antibody. J Clin Oncol. 2019;37(30):2730–7.

32. Eggermont AMM, Kicinski M, Blank CU, Mandala M, Long GV, Atkinson V, et al. Association Between Immune-Related Adverse Events and Recurrence-Free Survival Among Patients With Stage III Melanoma Randomized to Receive Pembrolizumab or Placebo: A Secondary Analysis of a Randomized Clinical Trial. JAMA Oncol. 2020;6(4):519–27.

33. Shankar B, Zhang J, Naqash AR, Forde PM, Feliciano JL, Marrone KA, et al. Multisystem Immune-Related Adverse Events Associated With Immune Checkpoint Inhibitors for Treatment of Non-Small Cell Lung Cancer. JAMA Oncol. 2020;6(12):1952–6.

34. Valero C, Lee M, Hoen D, Weiss K, Kelly DW, Adusumilli PS, et al. Pretreatment neutrophil-to-lymphocyte ratio and mutational burden as biomarkers of tumor response to immune checkpoint inhibitors. Nat Commun. 2021;12(1):729.

35. Wang Z, Aguilar EG, Luna JI, Dunai C, Khuat LT, Le CT, et al. Paradoxical effects of obesity on T cell function during tumor progression and PD-1 checkpoint blockade. Nat Med. 2019;25(1):141–51.

36. Dyck L, Prendeville H, Raverdeau M, Wilk MM, Loftus RM, Douglas A, et al. Suppressive effects of the obese tumor microenvironment on CD8 T cell infiltration and effector function. J Exp Med. 2022;219(3).

37. Pingili AK, Chaib M, Sipe LM, Miller EJ, Teng B, Sharma R, et al. Immune checkpoint blockade reprograms systemic immune landscape and tumor microenvironment in obesity-associated breast cancer. Cell Rep. 2021;35(12):109285.

38. Arteaga DP, DeKraker C, Ennis M, Dewey N, Goebel EA, Welch S, et al. Body composition and endometrial cancer outcomes. J Natl Cancer Inst Monogr. 2023;2023(61):49–55.

39. Celik E, Kizildag Yirgin I, Goksever Celik H, Engin G, Sozen H, Ak N, et al. Does visceral adiposity have an effect on the survival outcomes of the patients with endometrial cancer? J Obstet Gynaecol Res. 2021;47(2):560–9.

40. Moukarzel LA, Ferrando L, Stylianou A, Lobaugh S, Wu M, Nobre SP, et al. Impact of obesity and white adipose tissue inflammation on the omental microenvironment in endometrial cancer. Cancer. 2022;128(18):3297–309.

41. Jamieson A, Bosse T, and McAlpine JN. The emerging role of molecular pathology in directing the systemic treatment of endometrial cancer. Ther Adv Med Oncol. 2021;13:17588359211035959.

42. McAlpine J, Leon-Castillo A, and Bosse T. The rise of a novel classification system for endometrial carcinoma; integration of molecular subclasses. J Pathol. 2018;244(5):538–49.

43. Leon-Castillo A, de Boer SM, Powell ME, Mileshkin LR, Mackay HJ, Leary A, et al. Molecular Classification of the PORTEC-3 Trial for High-Risk Endometrial Cancer: Impact on Prognosis and Benefit From Adjuvant Therapy. J Clin Oncol. 2020;38(29):3388–97.

44. Oaknin A, Tinker AV, Gilbert L, Samouelian V, Mathews C, Brown J, et al. Clinical Activity and Safety of the Anti-Programmed Death 1 Monoclonal Antibody Dostarlimab for Patients With Recurrent or Advanced Mismatch Repair-Deficient Endometrial Cancer: A Nonrandomized Phase 1 Clinical Trial. JAMA Oncol. 2020;6(11):1766–72.

45. Fader AN, Roque DM, Siegel E, Buza N, Hui P, Abdelghany O, et al. Randomized Phase II Trial of Carboplatin-Paclitaxel Compared with Carboplatin-Paclitaxel-Trastuzumab in Advanced (Stage III-IV) or Recurrent Uterine Serous Carcinomas that Overexpress Her2/Neu (NCT01367002): Updated Overall Survival Analysis. Clin Cancer Res. 2020;26(15):3928–35.

46. Horeweg N, Workel HH, Loiero D, Church DN, Vermij L, Leon-Castillo A, et al. Tertiary lymphoid structures critical for prognosis in endometrial cancer patients. Nat Commun. 2022;13(1):1373.

47. Dai Y, Zhao L, Hua D, Cui L, Zhang X, Kang N, et al. Tumor immune microenvironment in endometrial cancer of different molecular subtypes: evidence from a retrospective observational study. Front Immunol. 2022;13:1035616.

48. Dessources K, Ferrando L, Zhou QC, Iasonos A, Abu-Rustum NR, Reis-Filho JS, et al. Impact of immune infiltration signatures on prognosis in endometrial carcinoma is dependent on the underlying molecular subtype. Gynecol Oncol. 2023;171:15–22.

49. Ramos-Casals M, Brahmer JR, Callahan MK, Flores-Chavez A, Keegan N, Khamashta MA, et al. Immune-related adverse events of checkpoint inhibitors. Nat Rev Dis Primers. 2020;6(1):38.

50. Banack HR, and Stokes A. The ‘obesity paradox’ may not be a paradox at all. Int J Obes (Lond). 2017;41(8):1162–3.

51. Eisenhauer EA, Therasse P, Bogaerts J, Schwartz LH, Sargent D, Ford R, et al. New response evaluation criteria in solid tumours: revised RECIST guideline (version 1.1). Eur J Cancer. 2009;45(2):228–47.

52. Momeni-Boroujeni A, Dahoud W, Vanderbilt CM, Chiang S, Murali R, Rios-Doria EV, et al. Clinicopathologic and Genomic Analysis of TP53-Mutated Endometrial Carcinomas. Clin Cancer Res. 2021;27(9):2613–23.

53. Leon-Castillo A, Britton H, McConechy MK, McAlpine JN, Nout R, Kommoss S, et al. Interpretation of somatic POLE mutations in endometrial carcinoma. J Pathol. 2020;250(3):323–35.

54. Middha S, Zhang L, Nafa K, Jayakumaran G, Wong D, Kim HR, et al. Reliable Pan-Cancer Microsatellite Instability Assessment by Using Targeted Next-Generation Sequencing Data. JCO Precis Oncol. 2017;2017.

